# Preliminary Non-Randomized Clinical Trial of Subcutaneous Esketamine in Treatment-Resistant Depression: Exploring Adjunctive Effects of Ketamine-Assisted Psychotherapy

**DOI:** 10.64898/2026.05.31.26354555

**Authors:** Yves Martins Varela, Patrícia Cavalcanti-Ribeiro, Geovan Menezes de Sousa, Marcelo Falchi-Carvalho, Juliana dos Santos Fernandes Barbalho, Raynara Bolcont de Oliveira Gomes, Melanie Moura Medina Gurgel, Bruno Carvalho Pereira, Pâmela Milena de Lima Souza, Kaike Thiê da Costa Gonçalves, Mariana Muniz, Victor Rocha Nóbrega de Almeida, Luiz Felipe Dantas Pereira, David Cavalcante Barbosa, Bruna Santos de Carvalho, Eduardo Igor Torquato Cardoso Lopes, Ariadne Cruz de Oliveira, Draulio Barros de Araujo, Fernanda Palhano-Fontes, Gisele Fernandes-Osterhold, Nicole Galvão-Coelho

## Abstract

**Background:** Ketamine has emerged as an effective rapid-acting treatment for treatment-resistant depression (TRD), producing significant antidepressant effects within hours of administration. Given ketamine’s capacity to induce states of heightened neuroplasticity and psychological openness, psychotherapy may represent a meaningful complement to its pharmacological effects — facilitating emotional processing, cognitive restructuring, and the consolidation of therapeutic gains. However, the adjunctive potential of structured psychotherapeutic support in ketamine-based interventions remains largely unexplored.

**Methods:** This preliminary, non-randomized, open-label clinical trial evaluated the adjunctive effects of ketamine-assisted psychotherapy (KAP) in an outpatient setting. Forty-six patients with TRD received eight weekly sessions of subcutaneous esketamine (0.5–1.0 mg/kg) and were allocated into two groups: esketamine without psychotherapeutic support (n = 23) and esketamine combined with structured KAP encompassing preparation, dosing accompaniment, and post-session integration (n = 23). Depressive symptoms were assessed using the Montgomery–Åsberg Depression Rating Scale (MADRS) and the Beck Depression Inventory-II (BDI-II) at multiple timepoints during treatment and at follow-up assessments up to six months after protocol completion.

**Results:** Both groups showed significant reductions in depressive symptoms throughout treatment. The KAP group demonstrated greater clinical improvement by the end of treatment, with between-group differences on the MADRS emerging at sessions 7 and 8. MADRS response and remission rates were 52.2% and 34.8% in the KET group, and 78.3% and 78.3% in the KAP group, respectively. BDI-II scores indicated earlier subjective improvement in the KAP group, with between-group differences emerging as early as the second session and persisting across multiple timepoints. No significant between-group differences were observed during the six-month follow-up, with both groups maintaining symptom reductions comparable to end-of-treatment levels.

**Conclusions:** These findings suggest that structured psychotherapeutic support may be associated with early clinical response and remission rates in subcutaneous esketamine treatment for TRD, potentially through facilitation of emotional processing, psychological flexibility, and behavioural change. Further controlled studies are needed to clarify the specific contribution of psychotherapy, investigate the mechanisms underlying this interaction, and optimize integrated treatment approaches for TRD. The trial was registered at https://ensaiosclinicos.gov.br/rg/RBR-1072m6nv.

## Introduction

Over the past two decades, ketamine has emerged as one of the most significant advances in the treatment of treatment-resistant depression (TRD), demonstrating rapid antidepressant effects within hours of administration. It is a non-competitive antagonist of N-methyl-D-aspartate (NMDA) glutamatergic receptors that promotes synaptogenesis through downstream activation of AMPA receptors and modulation of neuroplasticity-related signalling pathways, including BDNF-TrkB and mTOR (Krystal et al., 2024; Pribish et al., 2020). In controlled trials, response rates following repeated intravenous infusions have reached approximately 70% (Zarate et al., 2006; Murrough et al., 2013); however, real-world data tell a more conservative story. A systematic review and meta-analysis of 79 studies encompassing over 2,600 patients found mean response and remission rates of approximately 45% and 30%, respectively, with considerable variability across clinical populations (Alnefeesi et al., 2022). Furthermore, even among initial responders, the majority — approximately 70% — relapse within six months of treatment discontinuation (Zaki et al., 2025), underscoring the limited durability of pharmacological effects alone.

This gap between controlled efficacy and real-world effectiveness raises the question of whether adjunctive interventions, particularly psychotherapeutic support – including structured psychological support, therapeutic preparation, dosing accompaniment, and post-session integration – may enhance or prolong ketamine’s therapeutic effects. Some early studies had already begun to explore the therapeutic relevance of ketamine beyond its pharmacological effects alone. Fontana and Loschi (1974) were among the first to document its use as an adjunct to psychodynamic psychotherapy in depressed patients, while Krupitsky and Grinenko (1997) later demonstrated in a randomized trial that ketamine combined with psychedelic psychotherapy yielded substantially higher one-year abstinence rates in patients with alcohol dependence compared to conventional treatment (65.8% vs. 24%). Nevertheless, subsequent decades of research prioritized biomedical validation — dose-finding, mechanism of action, and route of administration — culminating in the regulatory approval of intranasal esketamine for TRD in 2019 (Food and Drug Administration, 2019) as a pharmacological intervention alone, without explicit consideration of adjunctive psychotherapeutic support, which remained largely unresearched.

Only recently has interest in combining ketamine with psychotherapy re-emerged as a research priority. This renewed attention reflects the growing recognition that ketamine alone often produces incomplete or short-lived responses in patients with TRD. A growing body of studies has since investigated whether adding structured psychotherapeutic support may enhance or prolong treatment outcomes. Systematic reviews suggest improvements not only in TRD, but also in anxiety, stress, pain, substance dependence, and depressive symptoms comorbid with other conditions (Dore et al., 2019; Drozdz et al., 2022; Varela et al., 2026). However, these studies remain highly heterogeneous regarding the psychotherapeutic approaches employed, the timing of psychotherapy relative to ketamine administration, and the duration of psychological interventions (Drozdz et al., 2022; Kew et al., 2023; Mallevays et al., 2026).

Within this context, ketamine-assisted psychotherapy (KAP) is a structured therapeutic model that goes beyond pharmacological administration by incorporating three phases: preparation, dosing support, and post-session integration (Dore et al., 2019; Mathai et al., 2022). Preparation sessions provide psychoeducation regarding ketamine’s effects and safety, orienting the patient to the range of possible experiences, and begin the building of the therapeutic alliance that will sustain engagement throughout the intervention (Dore et al., 2019; Drozdz et al., 2022). At subanesthetic doses, ketamine can elicit dissociative states, altered self-referential processing, and the spontaneous emergence of stressful or traumatic memories. While some researchers argue that these phenomena may actively facilitate therapeutic response and psychological change (Ballard and Zarate, 2020; Grabski et al., 2020; Mathai et al., 2020), they also carry the risk of traumatization when unaccompanied by adequate psychological support. Therapeutic accompaniment during dosing therefore serves not to suppress these states, but to help the patient navigate them safely within a contained relational context — transforming potentially destabilizing experiences into opportunities for meaningful psychological processing. Integration sessions, in turn, support the processing of cognitive, emotional, and relational material that emerges during dosing, and aim to consolidate new insights into meaningful shifts in attitudes, behaviours, and daily functioning (Dore et al., 2019; Muscat et al., 2021).

Not all KAP protocols, however, follow this three-phase structure. In practice, there is considerable variability in how psychotherapeutic support is operationalized across studies — ranging from fully structured models encompassing preparation, dosing accompaniment, and integration, to protocols in which psychotherapy is provided only before or after administration, or in which no structured support during dosing is offered at all (Varela et al., 2026). Beyond the presence or absence of specific phases, studies also differ in the number of sessions, the total duration of psychological interventions, the timing of psychotherapy relative to ketamine administration, the therapeutic modality employed, and the professional background of the therapist (Drozdz et al., 2022; Kew et al., 2023). Most protocols do not follow a specific psychotherapeutic school, instead adopting generalist or integrative frameworks combining techniques from different approaches (Cavarra et al., 2022; Nutt et al., 2025). This heterogeneity makes it difficult to compare findings across studies, and, to date, no consensus exists on an optimal KAP protocol (Drozdz et al., 2022).

To date, only one study has directly examined outcomes in patients receiving ketamine with versus without adjunctive psychotherapy for depression and PTSD (Moore et al., 2025). Using a naturalistic observational design with help-seeking samples, the study found no significant differences between conditions. However, its conclusions are substantially limited by the fact that it was not specifically designed to test the role of psychotherapeutic support: participants in the psychotherapy condition received heterogeneous interventions delivered by therapists with different backgrounds and theoretical orientations, the ketamine protocols were not standardized across participants, and crucially, no structured psychological support was provided during dosing sessions — an element considered both ethically and technically essential in psychedelic-assisted interventions (Luoma et al., 2022; Moore et al., 2025). These limitations underscore the need for studies employing a controlled and standardized ketamine protocol alongside structured psychotherapeutic support explicitly encompassing all three phases of KAP.

The present study addresses this gap by comparing a standardized subcutaneous esketamine protocol delivered as a conventional biomedical intervention with the same pharmacological protocol delivered within a structured KAP-informed care model, including preparation, dosing accompaniment, post-session integration, individualized setting, and continuous therapeutic support. Compared to intravenous delivery, SC administration offers ease of application, fewer acute cardiovascular effects, and plasma concentrations comparable to intramuscular administration (Chilukuri et al., 2014; Gálvez et al., 2014; George et al., 2017; Glue et al., 2011; Loo et al., 2016). Its lower cost relative to both intravenous and intranasal esketamine further increases its accessibility, particularly in real-world clinical settings with limited resources (Palhano-Fontes et al., 2024). Previous work from our group has demonstrated the efficacy and safety of repeated SC esketamine in TRD, including rapid and sustained antidepressant effects (Palhano-Fontes et al., 2024) and significant reductions in suicidality lasting up to six months (Lopes et al., 2024), providing a well-characterized pharmacological platform on which to investigate the contribution of adjunctive psychotherapeutic support.

To our knowledge, this is the first study specifically designed to compare subcutaneous esketamine with and without structured KAP — encompassing preparation, dosing accompaniment, and post-session integration with explicit psychoeducation and processing of dissociative experiences — within a standardized pharmacological protocol. We hypothesize that the addition of structured psychotherapeutic support will enhance clinical outcomes beyond pharmacological effects alone, reflected in higher response and remission rates and greater durability of antidepressant effects over follow-up.

## Materials and Methods

### Study design

This study consisted of an open-label, non-randomized clinical trial conducted in two sequential phases, designed to evaluate the adjunctive effects of structured psychotherapeutic support in patients undergoing subcutaneous esketamine treatment for treatment-resistant depression (TRD). In Phase 1, a convenience sample of patients received subcutaneous esketamine without psychotherapeutic support (KET group); clinical outcomes from this phase have been previously reported (Palhano-Fontes et al., 2024; Lopes et al., 2024). In Phase 2, a new convenience sample of patients received the same esketamine protocol combined with structured ketamine-assisted psychotherapy (KAP group), encompassing preparation, dosing accompaniment, and post-session integration sessions. The Phase 2 sample was recruited by convenience sampling, with a target sample size matched to Phase 1. Despite the absence of randomization, the two groups were comparable in key demographic and clinical characteristics at baseline, as reported in the results section. This study was conducted at the Hospital Universitário Onofre Lopes (HUOL/UFRN) and approved by the institution’s Research Ethics Committee under protocol number 39222920.2.0000.5292. The trial was registered at the Registro Brasileiro de Ensaios Clínicos (ReBEC) (https://ensaiosclinicos.gov.br/rg/RBR-1072m6nv) and conducted in accordance with the Declaration of Helsinki.

### Participants and recruitment

A total of 57 adult patients with a current episode of treatment-resistant depression were enrolled in the study across two sequential phases. Recruitment occurred through community advertisements via digital media, referrals from local psychiatrists, and referrals from the hospital’s outpatient depression clinic. Interested candidates completed an online screening form filled out by their referring psychiatrist, which provided initial clinical information including previous diagnoses, treatment history, and prior medication trials. Following screening, participants underwent a structured psychiatric evaluation conducted by the study team using validated diagnostic instruments to confirm the diagnosis of treatment-resistant depression, rule out bipolar disorder, and verify compliance with all inclusion and exclusion criteria. Sociodemographic and clinical history data were collected at baseline. Written informed consent was obtained prior to enrolment. This evaluation process was consistent across both study phases, ensuring diagnostic comparability between groups.

### Inclusion and exclusion criteria

The study was restricted to patients experiencing a current moderate-to-severe depressive episode (Montgomery–Åsberg Depression Rating Scale [MADRS] ≥ 25) lasting for at least four weeks and meeting criteria for treatment-resistant depression, defined as failure to achieve a satisfactory response following the use of at least two antidepressant medications from different pharmacological classes administered at adequate doses and duration (Caldioroli et al., 2021). Exclusion criteria included physical and mental health conditions that could interfere with the safe administration of esketamine, including uncontrolled hypertension, congestive heart failure or other evidence of cardiac impairment, chronic obstructive pulmonary disease, severe obesity, increased intracranial or cerebrospinal pressure, pregnancy,hyperthyroidism, previous adverse reactions to esketamine, current or past psychotic symptoms, substance use disorders, dissociative disorders, autism spectrum disorders, and prodromal symptoms of schizophrenia. Individuals deemed ineligible or who chose to withdraw from the study at any point returned to standard treatment.

### Instruments and data collection procedures

All instruments were administered consistently across both study phases.

Montgomery–Åsberg Depression Rating Scale (MADRS) (Montgomery and Åsberg, 1979):

The MADRS is a clinician-administered scale assessing the severity of depressive symptoms across ten items, yielding scores classified as normal (0–6), mild (7–19), moderate (20–34), or severe (>34). It was administered during screening and immediately before each treatment session. Item 10 of the scale was used to monitor suicidal ideation (MADRS-SI) throughout the study. Clinical response was defined as a reduction of 50% or more in MADRS scores relative to baseline, and remission as a score ≤ 12 at end of treatment.

Beck Depression Inventory-II (BDI-II) (Beck et al., 1996): The BDI-II is a 21-item self-report measure assessing depressive symptom severity, with scores categorized as minimal or absent (0–10), mild (11–19), moderate (20–30), and severe (31–63). It was administered at baseline, immediately before each dosing session, 24 hours after each session, and at follow-up assessments conducted at 1, 2, 4, and 6 months after treatment completion.

Clinician-Administered Dissociative States Scale (CADSS) (Bremner et al., 1998): The CADSS is a clinician-administered instrument designed to assess the presence and intensity of dissociative and altered states of consciousness. In the present study, it was administered 40 to 60 minutes after esketamine dosing in both groups to capture the peak period of ketamine-induced psychoactive effects. Full results from the CADSS will be reported in a subsequent secondary analysis.

Additional instruments were administered throughout the study, including the Beck Scale for Suicidal Ideation (BSI), the Beck Anxiety Inventory (BAI), the Pittsburgh Sleep Quality Index (PSQI), and the Positive and Negative Affect Schedule (PANAS). Results from these instruments will be reported in subsequent secondary analyses. The present report focuses on depressive symptom severity as assessed by the MADRS and BDI-II as primary outcome measures.

### Procedures

Participants in both groups underwent eight weekly sessions of subcutaneous esketamine administration following the same pharmacological protocol. The key difference between groups was the presence or absence of structured psychotherapeutic support: the KET group received esketamine administration without psychotherapy, whereas the KAP group received esketamine combined with structured ketamine-assisted psychotherapy encompassing two 90-minute preparatory sessions prior to treatment onset, a 30-minute preparation session immediately before each dosing session, and a 60-minute integration session conducted 24 hours afterward. Furthermore, the environmental conditions provided for each group aimed to reproduce the specificities of each setting during the dosage sessions: the KET group received medical supervision in an outpatient room (with capacity to accommodate up to two patients at a time); the KAP group had psychotherapeutic support from a doctor and a psychologist in a private environment decorated for psychotherapy sessions (indirect lighting, decoration with abstract art paintings and a wooden panel superimposed on the wall). To ensure the volunteers’ comfort, both groups had access to a fluffy blanket, blindfolds, headphones, and a playlist selected to facilitate immersion in the experience.

All participants were instructed to fast for four hours and abstain from liquid intake for two hours before each session, which typically began at 8:00 a.m. in the psychiatric unit of the Hospital Universitário Onofre Lopes. Blood samples were collected prior to each dosing session for biochemical assessments, including renal and hepatic function markers as well as inflammatory and stress biomarkers. Pre-dose assessments (MADRS and BDI-II) were conducted immediately before each dosing session and seven days after the last session.

Esketamine hydrochloride was administered subcutaneously in the lateral region of the umbilical scar using a dose-titration approach: the initial dose was 0.5 mg/kg, increasing to 0.75 mg/kg and subsequently to 1.0 mg/kg in sessions where adequate response — defined as ≥ 50% symptom improvement or remission — had not been achieved. During the first session, the dose was divided into two injections separated by a 15-minute interval; subsequent sessions were not divided unless adverse effects required adjustment. All participants maintained their usual medication regimens throughout the study; however, those using benzodiazepines were instructed to discontinue use 24 hours before and after each dosing session, given their potential to attenuate the psychoactive and dissociative effects of esketamine. Participants using Z-drugs (non-benzodiazepine hypnotics acting on GABA-A receptors, such as zolpidem and zopiclone) were also instructed to discontinue use during the same period, as a precautionary measure due to potential adverse interactions with esketamine.

During dosing sessions, all participants of both groups remained in a reclining chair listened to music through headphones and the vital signs — including heart rate, systolic and diastolic blood pressure, and oxygen saturation (SpO2) — were recorded every 15 minutes throughout each session in both groups. In the KET group, sessions were conducted in a clinical environment room accommodating two patients separated by a partition, with a nurse present exclusively for medical monitoring and management of potential adverse events, and the music was selected from available playlists without specific therapeutic design. In the KAP group, participants were seen individually in a private room designed to provide a comfortable, non-clinical environment, featuring indirect lighting, wallpaper, and décor intended to reduce the institutional character of the setting and support therapeutic engagement. They were continuously accompanied by both a psychiatrist and a psychologist, and a purpose-designed music program was used to support the therapeutic experience.

The Clinician-Administered Dissociative States Scale (CADSS) was administered 40 to 60 minutes after dosing in both groups to assess the intensity and quality of altered states of consciousness induced by esketamine. After 90 minutes, patients underwent clinical evaluation and were subsequently discharged. Prior to discharge, all participants were evaluated using the modified Aldrete score (Aldrete, 1995) to confirm clinical stability, and discharge was authorized only in the presence of a responsible companion. One day after each session, participants completed the BDI-II online. This same scale was applied online at follow-up phase: 1, 2, 4, and 6 months after treatment completion.

Cardiovascular parameters, renal and hepatic function data from Phase 1 have been previously reported (Palhano-Fontes et al., 2024), and anti-inflammatory effects of the subcutaneous esketamine protocol observed in Phase 1 are described in Cavalcanti-Ribeiro et al. (2025).

The study design is illustrated in Figure 1.

**Figure 1.**
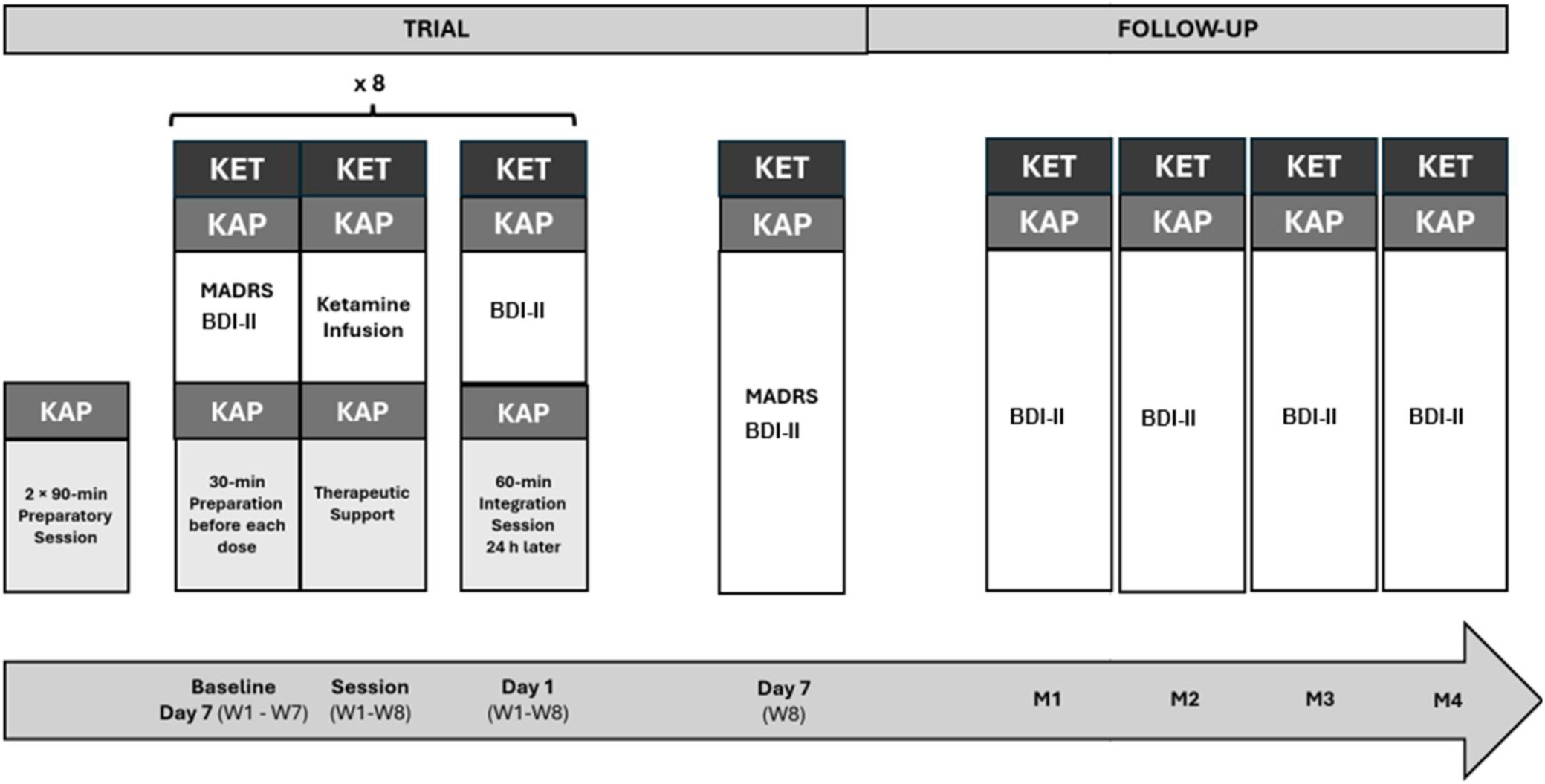
Study Design. *Note:* Participants underwent eight weekly esketamine dosing sessions assigned either to ketamine therapy without psychotherapy (KET) or ketamine-assisted psychotherapy (KAP). KET consisted solely of weekly esketamine administrations. KAP additionally included two 90-min preparatory sessions prior to the first dosing, a 30-min preparatory session before each weekly dose, in-session support, and a 60-min integration session 24 hours later. Pre-dose assessments (MADRS and BDI-II) were conducted immediately before each session (Week 1 – Week 8), and 7 days after the last session (Day 7 of week 8). Post-dose assessment (BDI-II) was completed online 24 h later each session and through follow-up phase: 1, 2, 4, and 6 months after the trial.

### Ketamine-assisted psychotherapy

In this study, the ketamine-assisted psychotherapy program consisted of three stages conducted by psychotherapists: preparation, dosing support, and integration. The first stage, preparation, was conducted by a psychologist and scheduled after participant inclusion in the study and before the first esketamine administration. This stage encompassed two 90-minute sessions one occurring up to one week before treatment initiation and the other occurring 48 to 72 hours before the first dosing session, aimed at establishing the therapeutic alliance, reducing treatment-related anxiety, and preparing the participant for the substance experience. During these sessions, participants received psychoeducation regarding depression and the potential dissociative and altered states of consciousness that could emerge during administration, with the goal of fostering familiarity, realistic expectations, and a sense of safety prior to the first dosing session.

Preparatory sessions also included a brief and non-directive exploration of the participant’s life history and possible traumatic experiences. Deliberate deep exploration of traumatic content was intentionally avoided at this stage, as it was understood that such material would naturally emerge and could be therapeutically processed throughout subsequent integration sessions. Emotional awareness, experiential acceptance, and an attitude of openness and curiosity toward the experiences that might arise during dosing sessions were encouraged as orienting dispositions throughout the treatment process. Mindfulness and emotional regulation practices were introduced and practiced as tools to support navigation of the psychedelic experience, and participants’ questions and concerns regarding the procedure were addressed.

The second stage, dosing support, consisted of two-hour sessions structured in two sequential parts, participants were continuously accompanied by a pair of therapists, a psychologist and a psychiatrist, trained to manage physical effects, dissociative experiences, and emotional and somatic dysregulation that may arise during altered states. The first 30 minutes comprised an immediate preparation period conducted by the psychologist, during which therapeutic agreements were reviewed, intentions for the session were established, participants’ questions and concerns were addressed, and grounding or mindfulness techniques were applied when indicated. Esketamine administration then commenced, with the dosing and therapeutic support phase lasting approximately 90 minutes. During esketamine effects, therapists maintained a non-directive and supportive presence throughout, encouraging participants to adopt an attitude of openness and curiosity and an observer perspective toward their experiences. Verbalization was encouraged only when participants considered it necessary. When clinically indicated, therapists employed specific techniques within the protocol framework, including mindfulness practices, grounding techniques, guided breathing, and therapeutic touch when explicitly requested by the participant — all aimed at providing sufficient containment for the processing of distressing material while minimizing the risk of traumatization. Each dosing session lasted approximately two hours in total.

The third stage, integration, was conducted via secure videoconference to protect participant privacy, on the day following each esketamine administration and one week after the final dosing session, totalling nine 60-minute sessions. Integration sessions were scheduled within 24 hours after dosing to take advantage of the period of heightened neuroplasticity that follows ketamine administration — a window during which mTOR-mediated synaptogenesis and increased synaptic plasticity in the prefrontal cortex may facilitate the consolidation of therapeutically relevant insights (Li et al., 2010; Krystal et al., 2024). The primary objectives of this phase were to provide a supportive space for empathic processing of the dosing experience, to assist participants in managing concerns, emotional, and somatic reactions arising during sessions, and to collaboratively construct meaning from the experience in relation to the participant’s life history. Integration sessions thus aimed not only at processing what occurred during dosing, but at consolidating experiential learning into sustainable change beyond the therapeutic context.

Across the full KAP protocol, each participant received approximately 27 hours of structured psychotherapeutic support: three hours of preparation, 16 hours of dosing accompaniment, and eight hours of integration sessions.

The protocol was adapted from the literature on psychedelic-assisted therapy (Dore et al., 2019; Winkelman and Sessa, 2019; Coleman, 2017), somatic approaches (Porges, 2011; Levine, 1999, 2012; Ross, 2014), cognitive approaches for trauma treatment (Resick et al., 2016; Shapiro, 2017), and contextual cognitive therapies (Hayes et al., 2011). It follows a methodological trend commonly observed in clinical trials involving psychedelic-assisted psychotherapy, namely the use of integrative frameworks not restricted to a single therapeutic school (Cavarra et al., 2022; Leone et al., 2024; Nutt et al., 2025).

A total of six psychologists participated in the study, with primary therapeutic orientations including Cognitive Behavioral Therapy (n = 3), Transpersonal Psychology (n = 2), and Psychoanalysis (n = 1). Although psychiatrists did not participate in preparation or integration sessions, their training in the KAP protocol was considered essential, as they accompanied each dosing session in full — from the immediate pre-dose preparation through to patient discharge. Therapists worked in pairs, a psychologist and a psychiatrist, the pairs were deliberately composed of members of different genders, in accordance with recommendations for psychedelic-assisted therapy settings that highlight the importance of gender-diverse therapeutic dyads as a safeguard for participant safety and comfort during states of heightened vulnerability and altered consciousness.

All therapists, as well as the medical team, underwent prior training and supervision provided by a collaborating psychologist affiliated with the University of California, San Francisco (UCSF), coordinator of facilitation in the psychedelic research program. Supervision encompassed synchronous and asynchronous instruction on psychedelic therapies and KAP management, case discussion meetings, intervention planning, educational and support materials, and guidance for the development of the present protocol. Supervision continued throughout the treatment period, providing ongoing support for clinical decision-making and protocol adherence.

### Statistical analysis

Depressive symptoms both assessed by a psychiatrist (MADRS) and self-reported (BDI-II evaluated 1-day [BDI-II_1_] and 7 days [BDI-II_7_] post-session) were used as dependent variables. The treatment group (KET or KAP) was used as the independent variable, and sessions as the repeated measures variable. Psychotherapy outside the study context was used as a covariate. Data analysis followed an intention-to-treat strategy, including all patients who completed assessments at the eight dosing sessions. To test the effect of treatments on dependent variables across sessions, a generalised linear model was performed with Fisher’s post-hoc tests, and p-values corrected for multiple comparisons. The model was based on a normal probability distribution with an identity link function, utilizing a robust covariance estimator (as suggested for small samples) and comparisons based on the Wald chi-square (*χ*^2^).

Response and remission proportions across sessions were compared between groups using a chi-square test with Yate’s correction for continuity. A repeated measure two-way ANOVA was used to evaluate vital sign changes across five time points (0, +15, +30, +45, and +60 minutes) and biochemical changes across three sessions (1st, 4th, and 8th), with group as the between factor and time points/session as the within factor. A multi-state Cox proportional hazard model was used to investigate whether dose evolution (0.5 → 0.75 → 1.0 mg/kg) was different for groups. Cox analysis was performed using R (4.5.0) and all other analyses were performed using SPSS (IBM).

## Results

### Sociodemographic and clinical characteristics

In the KET group, 82 individuals were screened, of whom 30 met eligibility criteria and later, seven participants discontinued the intervention: two participants were excluded due to a COVID-19 diagnosis prior to study initiation; one participant with pre-existing dystonia experienced symptom worsening and discontinued after the third session; one participant required hospitalization due to suicidal ideation and discontinued after the third session; one participant withdrew because of financial difficulties after the fourth session; and two participants were diagnosed with bipolar disorder during follow-up.

In the KAP group, 53 individuals were screened for eligibility, and 27 met inclusion criteria. Four participants withdrew for specific reasons that prevented continued attendance at weekly dosing sessions (two withdrew after the third session, two after the fourth session and one after the first session): one experienced a worsening of their clinical condition and required hospitalization; two did not tolerate the dissociative effects of esketamine and requested voluntary withdrawal from the study; and one developed hypomanic symptoms during the study, raising suspicion of bipolar disorder, resulting in withdrawal based on exclusion criteria guided by a protective clinical decision.

The final sample consisted of 23 Brazilian patients who received esketamine monotherapy (KET) and 23 patients who received esketamine treatment combined with psychotherapy (KAP). The CONSORT flowchart is presented in Figure 1.

**Figure 2.**
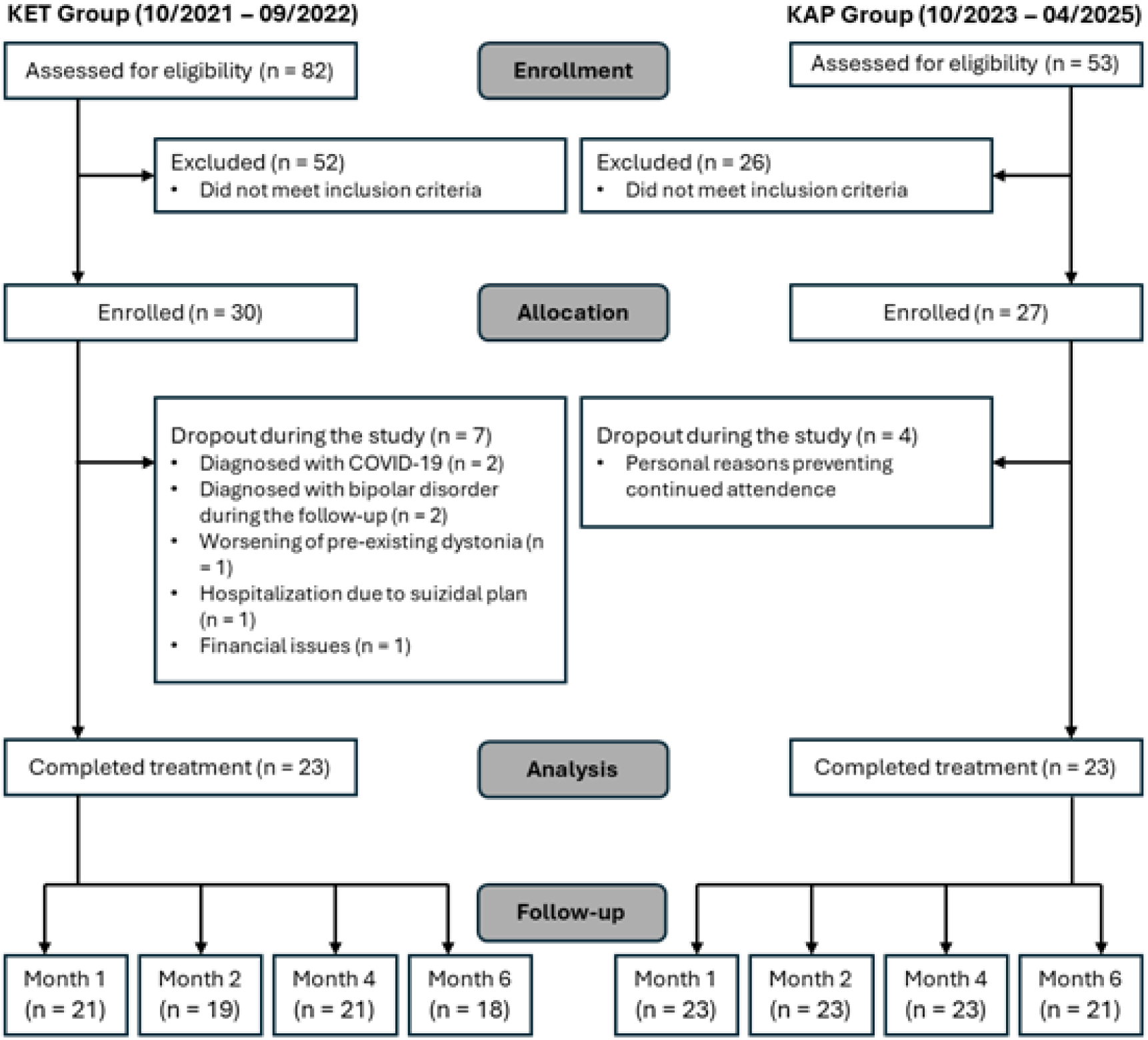
CONSORT flow chart. KET = ketamine monotherapy; KAP = ketamine-assisted psychotherapy.

A total of 46 patients received treatment: 23 received ketamine (Ketamine: 10/2021 – 09/2022) and 23, ketamine-assisted psychotherapy (KAP: 10/2023 – 04/2025). Most of the participants were women (Ketamine: n = 15, 65.2%; KAP: n = 12, 52.2%) with a mean age of 34.7 (SD = 12.7) years old for the KET group and 36.9 (SD = 11.5) years old for the KAP group. Most of the participants were single (Ketamine: 65.2%, n = 15; KAP: 65.2%, n = 15). Regarding education, the KET group showed a higher proportion of participants with 9 to 16 years of schooling (78.2%), whereas the KAP group was predominantly composed of participants with more than 16 years of education (60.9%). In terms of income, the KET group had a more balanced distribution across income levels, with 34.8% reporting high income (>10 minimum wages), compared to only 13.0% in the KAP group, which showed a predominance of moderate income (65.2%, 3–10 minimum wages). Regarding employment, the KET group had a lower proportion of employed participants (34.8%) compared to the KAP group (56.5%). Marital status was similar across groups, with most participants being single in both conditions (65.2% each). Engagement in physical activity outside the study was comparable between groups, less than the half practise regular physical activity (KET: 30.4%; KAP: 34.8%). Most participants attended to psychotherapy outside the context of the study, with a higher proportion of KET participants reported concurrent psychotherapy outside the study (82.6%) compared to the KAP group (60.9%); this variable was included as a covariate in subsequent analyses.

At baseline, both groups presented with moderate-to-severe depressive symptoms, as indicated by mean MADRS scores in the moderate range (KET: M = 30.9, SD = 4.9; KAP: M = 28.4, SD = 6.1) and severe self-reported depressive symptoms on the BDI-II (KET: M = 37.7, SD = 8.8; KAP: M = 35.7, SD = 11.7). Although overall severity was comparable, the clinical expression of chronicity differed between groups: the KET group showed a higher burden of recurrent episodes, with 30.4% reporting more than four depressive episodes, whereas the KAP group was characterized by longer episode duration, with a mean of approximately 10 years compared to 8 years in the KET group. Psychiatric comorbidities were common in both groups, with generalized anxiety disorder being the most frequently reported condition (KET: 60.9%; KAP: 52.2%). Most participants were using antidepressants at baseline (KET: 82.6%; KAP: 78.2%), and benzodiazepine use was reported by 5 participants in the KET group and 9 in the KAP group.

Sociodemographic and clinical characteristics of both groups are presented in Table 1. Between-group comparisons for these variables were conducted and are reported in the table.

**Table 1.** Sociodemographic and clinical characteristics.

|  | **KET (n = 23)** | **KAP (n = 23)** | **$\chi^2/t$ (p-value)** |
| --- | --- | --- | --- |
| Age (years) | 34.7 (12.7) | 36.9 (11.5) | -0.62 (0.538) |
| Sex |  |  |  |
| *Women* | 15 (65.2%) | 12 (52.2%) | 0.81 (0.369) |
| *Men* | 8 (34.8%) | 11 (47.8%) |  |
| Civil state |  |  |  |
| *Single* | 15 (65.2%) | 15 (65.2%) | 1.33 (0.513) |
| *Married* | 5 (21.7%) | 7 (30.4%) |  |
| *Divorced* | 3 (13.0%) | 1 (4.3%) |  |
| Education |  |  |  |
| *High school* | 1 (4.3%) | 5 (21.7%) | 10.77 (0.005) |
| *Under graduation* | 18 (78.2%) | 7 (30.4%) |  |
| *Graduation* | 4 (17.4%) | 11 (47.8%) |  |
| Employment |  |  |  |
| *Employed* | 8 (34.8%) | 13 (56.5%) | 1.40 (0.236) |
| *Unemployed* | 15 (65.2%) | 10 (43.5%) |  |
| Income (number of minimal wage) |  |  |  |
| *Low ($\leq 2$)* | 6 (26.1%) | 5 (21.7%) | 3.86 (0.145) |
| *Moderate (3-10)* | 9 (39.1%) | 15 (65.2%) |  |
| *High ($> 10$)* | 8 (34.8%) | 3 (13.0%) |  |
| Physical activity<br>(outside the study) | 7 (30.4%) | 8 (34.8%) | 0.00 (1.000) |
| Psychotherapy<br>(outside the study) | 19 (82.6%) | 14 (60.9%) | 2.68 (0.102) |
| MADRS | 30.9 (4.9) | 28.4 (6.1) | -0.37 (0.717) |
| Duration of depressive episode (years) | 7.96 (5.26) | 13.2 (9.63) | -2.30 (0.028) |
| Number of depressive episodes |  |  |  |
| 1 | 3 (13.0%) | 5 (21.7%) | 0.41 (0.815) |
| 2-4 | 11 (47.8%) | 11 (47.8%) |  |
| > 4 | 7 (30.4%) | 7 (30.4%) |  |
| Unknown | 2 (8.7%) | - |  |
| BDI-II | 37.7 (8.8) | 35.7 (11.7) | 2.02 (0.526) |
| Psychiatric comorbidities |  |  |  |
| PTSD | 1 (4.3%) | 0% (0) | 1.02 (0.312) |
| Panic disorder | 4 (17.4%) | 4 (17.4%) | 0.00 (1.000) |
| GAD | 14 (60.9%) | 52.2% (12) | 0.35 (0.552) |
| Social phobia | 5 (21.7%) | 3 (13.0%) | 0.605 (0.437) |
| Borderline Affective Disorder | 1 (4.3%) | 0 (0%) | 0.00 (1.000) |
| ADHD | 0 (0%) | 1 (4.3%) | 0.00 (1.000) |
| Use of medication |  |  |  |
| Antidepressant | 19 (82.6%) | 18 (78.2%) | 0.00 (1.000) |
| Mood stabiliser | 7 (30.4%) | 7 (30.4%) | 0.00 (1.000) |
| Atypical psychotics | 5 (21.7%) | 8 (34.8%) | 0.43 (0.513) |
| Typical psychotics | 0 (0%) | 1 (4.3%) | 0.00 (1.000) |
| Benzodiazepines | 5 (21.7%) | 9 (39.1%) | 0.92 (0.336) |
| Non-psychiatric medication | 5 (21.7%) | 5 (21.7%) | 0.00 (1.000) |

BDI-II = Beck Depression Inventory, MADRS = Montgomery-Åsberg Depression Rating Scale, ADHD = attention deficit and hyperactivity disorder, GAD = generalised anxiety disorder, PTSD = posttraumatic stress disorder. KET = ketamine monotherapy; KAP = ketamine-assisted psychotherapy.

### Esketamine safety and tolerability

The Cox regression model indicated that dose evolution was similar for both groups (HR = 0.90, CI 95%: 0.67 – 1.22, p = 0.507) (Figure S2). Regarding safety parameters, no group*session interaction was found for biochemical changes in urea, creatinine, ALT, AST, Na^+^, and K^+^ levels throughout the treatment, indicating that both groups showed similar and safe biochemical response to esketamine. Mean values for the assessed time points for both groups were within the populational reference values (Table S1).

Vital signs changed throughout the first hour of each dosing session (Figure 3, Tables S2 and S3). Both groups showed comparable baseline values for systolic blood pressure, diastolic blood pressure, and heart rate, except for oxygen saturation, which was significantly higher in the KAP group at baseline. Significant time effects were observed for systolic blood pressure (F_(4,1388)_ = 123.94, p < 0.001) and diastolic blood pressure (F_(4,1388)_ = 94.58, p < 0.001). Both measures increased significantly at +15 minutes in both groups. Systolic blood pressure remained elevated above baseline for the full 60 minutes in the KET group, returning to baseline by +45 minutes in the KAP group. For diastolic blood pressure, the KAP group showed significantly lower values than the KET group from +30 to +60 minutes (F_(4,1388)_ = 3.66, p = 0.006), also returning to baseline earlier than the KET group.

**Figure 3.**
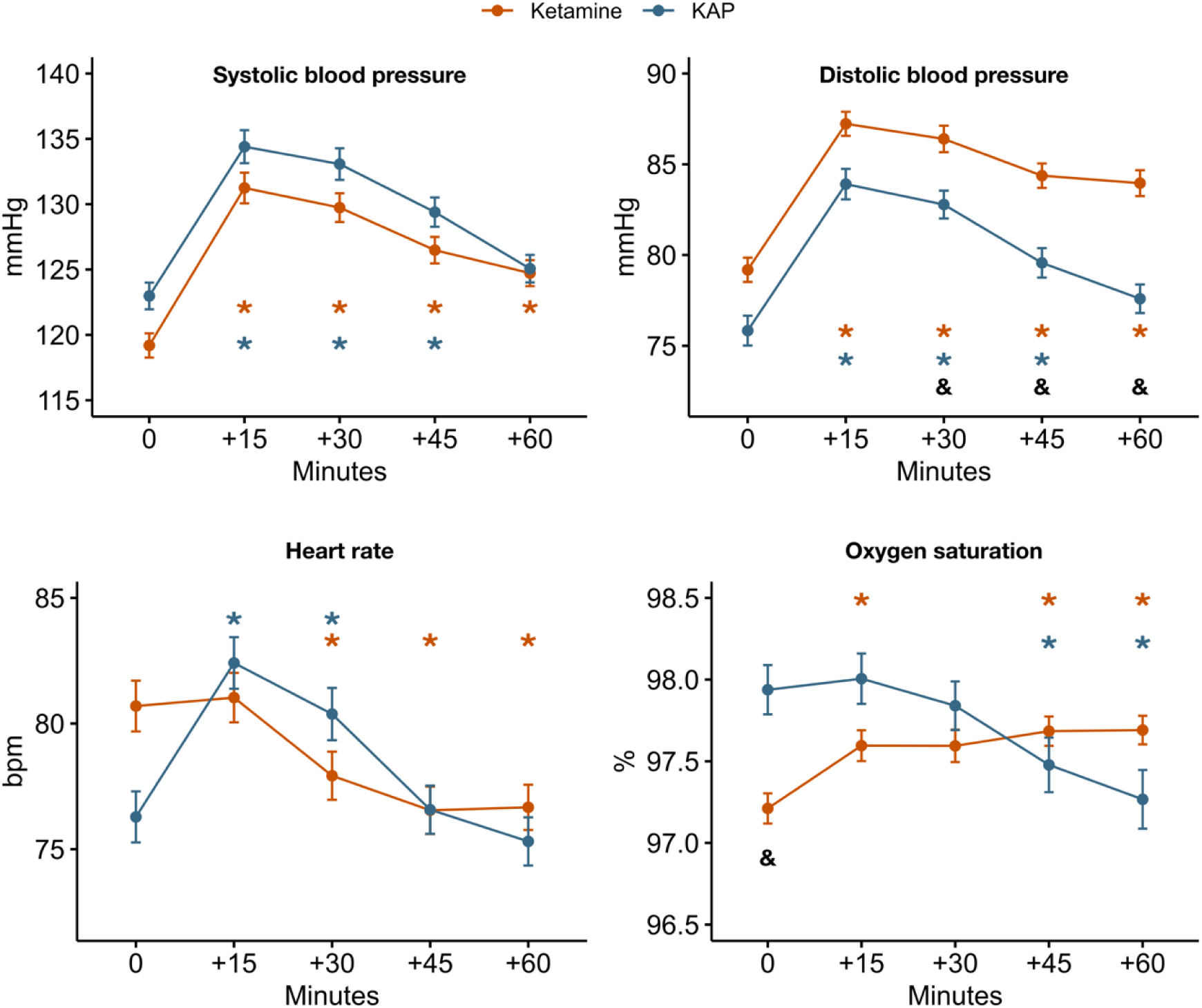
Changes in vital signs during acute (+15, +30, +45 and +60 minutes post-infusion) esketamine effects for KET and KAP groups. The coloured asterisks (*) indicate within-group difference with respect to baseline for both groups; the ampersand (&) indicates significant between-group difference. KET = ketamine monotherapy; KAP = ketamine-assisted psychotherapy.

Heart rate showed a significant time effect (F_(4,1384)_ = 53.05, p < 0.001), with distinct patterns between groups. The KET group showed a progressive reduction from +30 to +60 minutes post-infusion, while the KAP group showed increases at +15 and +30 minutes, returning to and remaining at baseline levels by +45 minutes.

Oxygen saturation also showed a significant time effect (F_(4,1276)_ = 15.67, p < 0.001). In the KAP group, SpO2 decreased significantly at +45 and +60 minutes relative to baseline, whereas in the KET group, SpO2 increased at +15, +45, and +60 minutes post-infusion. Values remained within clinically acceptable limits throughout the session in both groups, and no adverse cardiovascular events were recorded.

### Depressive symptoms – clinical assessment (MADRS)

There was a significant interaction between group and session (χ²(9)= 18,68, p = 0.028). At baseline, both groups showed comparable depressive symptom scores Even though both groups progressively decreased MADRS score throughout sessions (Figure 4, Table S4), KAP showed lower MADRS score at the end of treatment than the KET group (Session 7: p = 0.015; Session 8: p = 0.041) (Table S5). Psychotherapy outside the study was not a significant covariate (*χ*^2^(1) = 0.04, p = 0.851).

**Figure 4.**
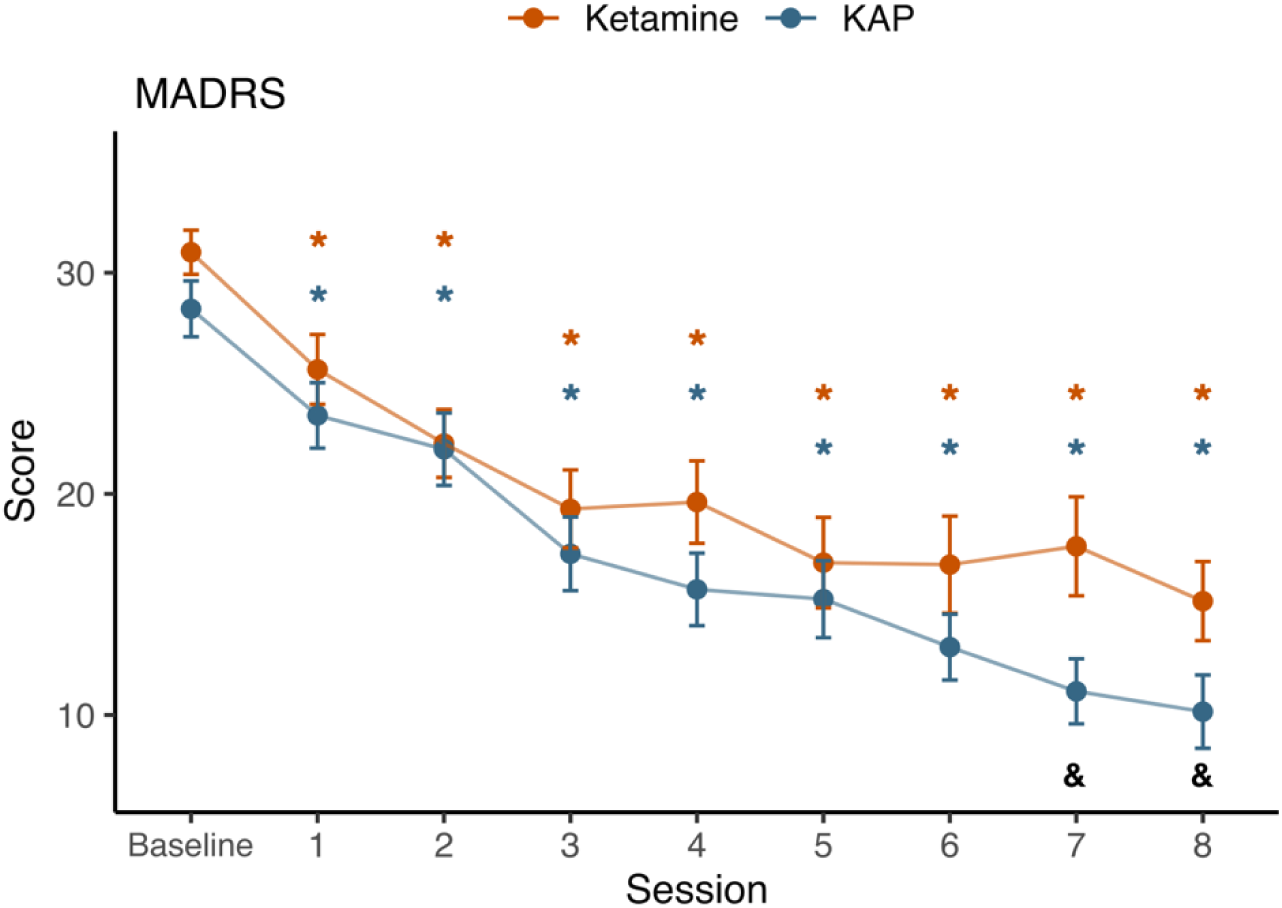
Mean depressive symptoms estimated by clinical assessment (MADRS scores) throughout the treatment sessions for the KET and KAP groups. The coloured asterisks (*) indicate differences detected in depressive symptoms relative to their baseline values for both groups in each session; the ampersand (&) indicates significant between-group difference. KET = ketamine monotherapy; KAP = ketamine-assisted psychotherapy.

Seven days after the first session, the response rate was 8.7% for the Ketamine group and 13% for the KAP group, while the remission rate was 4.3% and 8.7%, respectively. For KET, response rates increased until plateauing at 52.2% after the sixth session, remaining stable until the end of treatment, whereas in the KAP group, the rate continued to rise, reaching a maximum of 78.3% by the end of the treatment. However, no significant group differences were found (Table S4). The remission rate was similar between groups until the fourth session, when then the KAP group surpassed the KET group, which plateaued at 39.1%, although no significant difference was found (Table S4). By the end of treatment, remission rates were significantly (*χ*^2^ = 7.17, p = 0.007) higher for the KAP group (78.3%) than for the Ketamine group (34.8%) (Supplementary Table S4, Figure 5).

**Figure 5.**
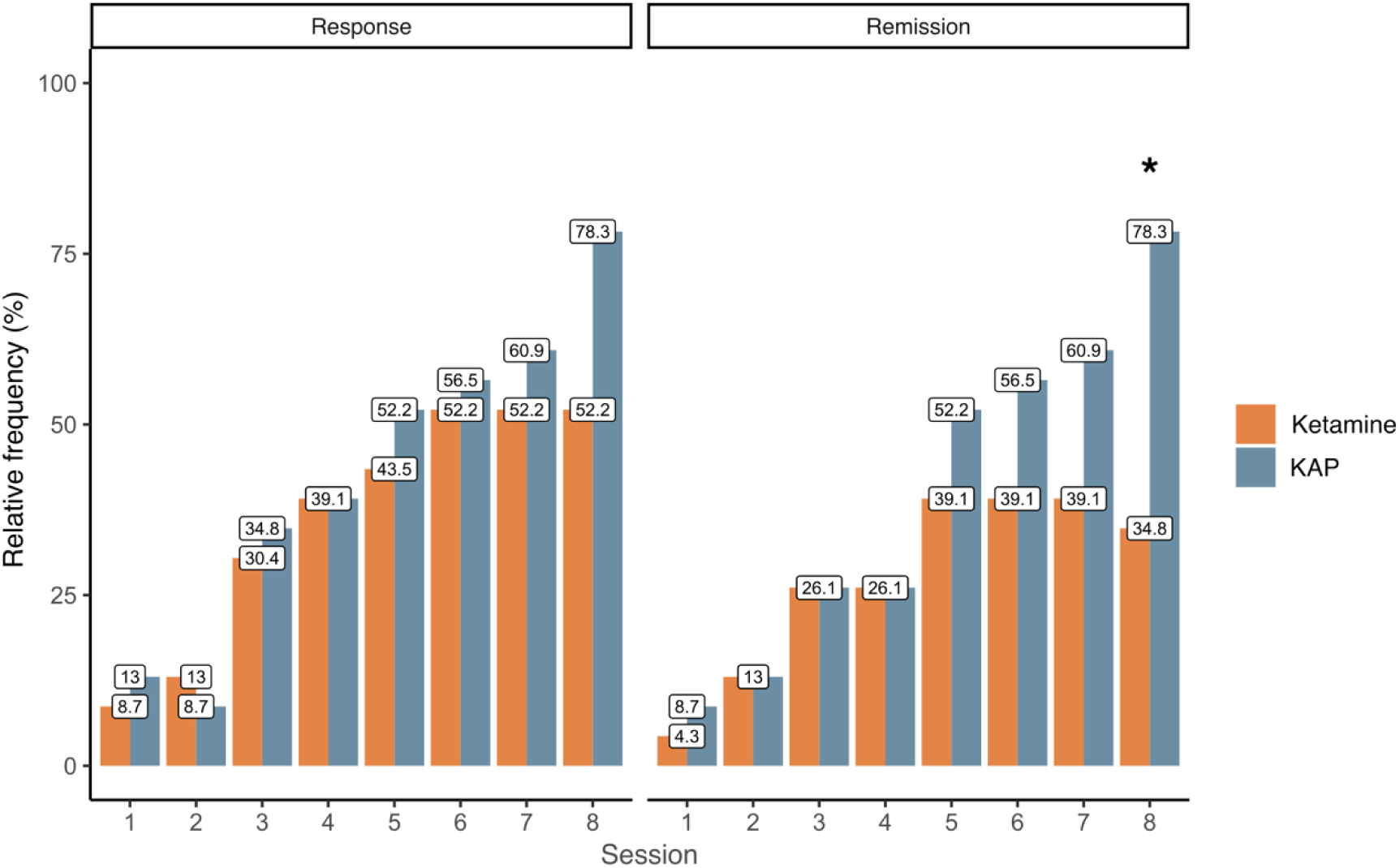
Response (left) and remission (right) rates for the KET and KAP groups. The rates are similar until Session 4, when the KAP group surpasses the KET group, particularly in the remission rates. KET = ketamine monotherapy; KAP = ketamine-assisted psychotherapy. * = between-group difference.

### Depressive symptoms – patients self-report (BDI-II)

At baseline, both groups showed comparable self-reported depressive symptom scores on the BDI-II, with no significant between-group difference (KET: M = 37.7, SD = 8.8; KAP: M = 35.7, SD = 11.7). Self-reported depressive symptoms assessed one day after each esketamine administration (BDI-II1) showed a significant group × session interaction (χ²(9) = 27.92, p = 0.001). Both groups demonstrated a progressive reduction in depressive symptoms throughout the treatment protocol relative to baseline (Figure 5A, Table S5). However, the KAP group showed significantly lower BDI-II scores than the KET group at sessions 2 (p = 0.030) and 3 (p = 0.026), with no significant between-group differences observed at the remaining sessions. (Figure 5B).

Depressive symptoms assessed seven days after each administration (BDI-II7) also showed a significant group × session interaction (χ²(9) = 33.86, p < 0.001). Both groups showed progressive symptom reduction throughout treatment relative to baseline, with the KAP group presenting significantly lower scores than the KET group at sessions 3 (p = 0.029), 4 (p = 0.006), 6 (p = 0.036), and 7 (p = 0.009), and no significant between-group differences at the remaining sessions (Figure 5A, Table S6).

When the two collection timepoints were compared within each group separately — examining whether scores differed between Day 1 and Day 7 post-session — neither group showed a significant main effect of collection timepoint (KET: χ²(1) = 0.82, p = 0.365; KAP: χ²(1) = 0.32, p = 0.570) nor a significant interaction between collection timepoint and session (KET: χ²(8) = 1.20, p = 0.997; KAP: χ²(8) = 1.21, p = 0.997) (Figure 5B, Table S6). This indicates that the reduction in self-reported depressive symptoms observed one day after each session was sustained throughout the following seven days for both groups.

Psychotherapy engagement outside the study sessions had no significant effect on BDI-II scores at either timepoint (BDI-II1: χ²(1) = 2.40, p = 0.122; BDI-II7: χ²(1) = 1.93, p = 0.165).

The follow-up analysis indicated that no differences were found between or within groups (χ²(5) = 2.01, p=0.848) (Figure 5C, Table S7) during the follow-up months relative to Session 8, suggesting that the effects found at the end of treatment were sustained similarly for both groups.

**Figure 5.**
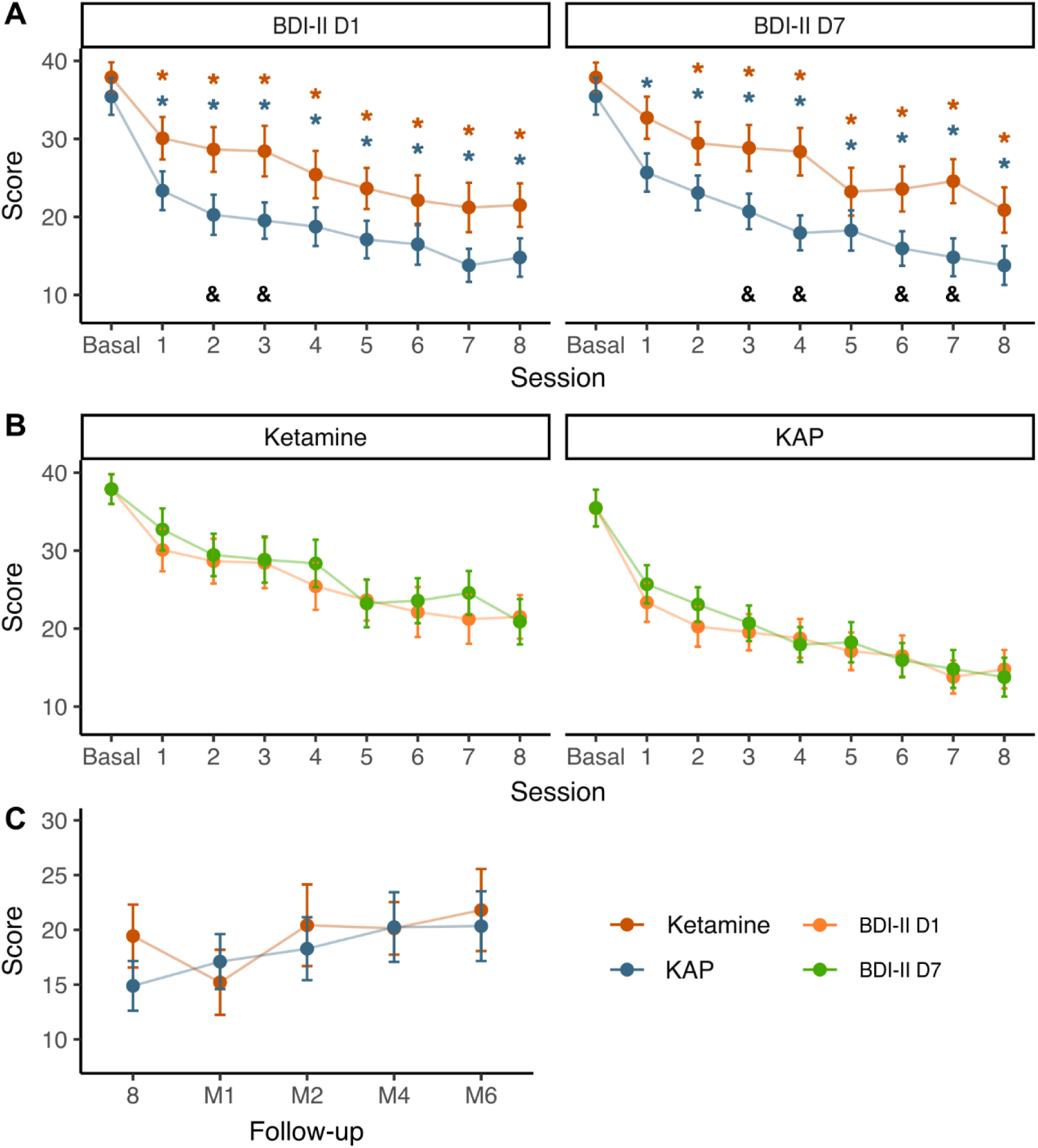
**A)** Mean depressive symptoms estimated by self-report (BDI-II scores, measured 1 day, left, and 7 days, right, post-treatment) throughout the treatment sessions for the Ketamine and KAP groups. **B)** BDI-II_1_ (orange) and BDI-II_7_ (green) measurements by group, highlighting the proximity of the measures and suggesting the sustenance of the effect obtained 1-day post-session until the following session. **C)** BDI-II follow-up measurements, showing that both groups sustain the responses obtained at the end of treatment. The coloured asterisks (*) indicate differences detected in depressive symptoms relative to their baseline values for both groups in each session; the black ampersand (&) indicates significant differences between groups.

## Discussion

The present study provides preliminary evidence for an adjunctive effect of structured psychotherapeutic support in the context of subcutaneous esketamine treatment for TRD. The KAP group demonstrated superior outcomes relative to the ketamine-only condition across both clinician-rated and patient-reported measures of depressive symptom severity although this between-group advantage was observed in the context of a sequential, open-label, non-randomized design and a multidimensional intervention contrast. Specifically, between-group differences were observed on the MADRS at sessions 7 and 8, and at multiple timepoints throughout treatment on the BDI-II, assessed both one and seven days post-session. Response and remission rates were also higher in the KAP group. Notably, the antidepressant effects observed within the first 24 hours after each session were sustained throughout the following seven days in both groups, suggesting a durable within-week effect of esketamine regardless of psychotherapeutic support — a finding consistent with previous results from our group using the same subcutaneous esketamine protocol (Palhano-Fontes et al., 2024).

However, contrary to our hypothesis, the adjunctive benefit of KAP did not appear to persist beyond the active treatment period. During follow-up, depressive symptoms remained reduced relative to baseline in both groups, at levels comparable to end-of-treatment scores, with no significant between-group differences observed over subsequent months. This pattern suggests that while structured psychotherapeutic support enhanced the antidepressant response during the treatment protocol, its long-term advantage over esketamine monotherapy was not maintained in the months following treatment completion.

Response and remission rates observed in Phase 1 — 52.2% and 34.8%, respectively — were consistent with those reported in the literature for multiple-dose ketamine protocols in TRD. For reference, Loo et al. (2023) reported response and remission rates of 29% and 19.6% respectively in the largest randomized controlled trial of subcutaneous ketamine to date, employing flexible-dose titration over 8 sessions in 4 weeks — the protocol most comparable to Phase 1 of the present study in terms of route and number of administrations. Higher rates have been reported with intravenous protocols employing greater numbers of infusions: Siegel et al. (2021) observed response and remission rates of 50% and 20% after 6 IV infusions, rising to 72% and 38% after 10 infusions in patients who continued beyond the initial induction phase — suggesting that the number of administrations may be an important determinant of clinical outcomes beyond route of administration alone. Phase 1 results from the present study fall within this range, supporting the clinical viability of the weekly SC protocol.

In contrast, Phase 2 yielded substantially higher rates for both response and remission (78.3% each), representing outcomes that exceed those typically reported for ketamine-based treatment without structured psychotherapeutic support, regardless of route or number of administrations. Notably, these rates also surpass those reported in the largest randomized controlled trials of intranasal esketamine combined with oral antidepressants, including the TRANSFORM and ESCAPE-TRD studies, which have reported remission rates in the range of 32–50% (Popova et al., 2019; Reif et al., 2023). To our knowledge, only one other study has reported comparable remission rates — a small retrospective case series of intranasal esketamine in Spain (N=11), in which 90.9% of patients achieved remission (Hernández et al., 2022). However, that study was limited by its very small sample, absence of a control group, retrospective design, and lack of a structured psychotherapeutic protocol, precluding meaningful comparison. The remission rates observed in Phase 2 of the present study, while notably high, must also be interpreted with caution given the relatively small sample size and non-randomized design. Nevertheless, these findings point to the potential clinical significance of structured psychotherapeutic support in enhancing remission outcomes in TRD and warrant replication in larger, randomized trials.

The present findings corroborate the therapeutic effects of ketamine-based interventions for TRD, consistent with results reported in systematic reviews of KAP studies (Drozdz et al., 2022; Kew et al., 2023). However, it is important to note that these reviews aggregated outcomes from studies in which all participants received some form of psychotherapeutic support alongside ketamine, without including a ketamine-only comparison condition. As such, they were not designed to isolate the adjunctive contribution of psychotherapy to treatment outcomes. To our knowledge, only one prior study has directly compared ketamine with and without adjunctive psychotherapy (Moore et al., 2025); however, its observational design, heterogeneous therapeutic approaches, and absence of structured support during dosing sessions substantially limit the conclusions that can be drawn. The present study addresses these limitations by employing a standardized pharmacological protocol and a structured three-phase KAP model, providing the first clinical trial specifically designed to evaluate the adjunctive effect of psychotherapeutic support in ketamine-based treatment for TRD.

The adjunctive effects of KAP emerged at different timepoints depending on the assessment perspective. From the clinician’s standpoint, as measured by the MADRS, between-group differences were observed only during the final two weeks of treatment — after the seventh esketamine administration — by which point both groups had already demonstrated clinical response. From the patient’s perspective, as measured by the BDI-II, adjunctive effects emerged earlier and more consistently, with between-group differences appearing as early as the second session and persisting across multiple timepoints throughout treatment. This temporal pattern is consistent with the known neurobiological profile of ketamine, which produces rapid and sustained antidepressant effects within the first hours and days of administration (Wu et al., 2021), potentially predominating over the more gradually emerging effects of psychotherapy in the early weeks of treatment. Indeed, evidence-based psychotherapies for depression typically require at least four to eight weeks to produce clinically significant improvements, even in non-resistant populations (American Psychological Association, 2006; Beck, 2021; Leichsenring and Steinert, 2017), making the emergence of adjunctive psychotherapeutic effects within approximately two months in a treatment-resistant sample a clinically meaningful finding. This temporal dynamic may reflect a bidirectional potentiation: while psychotherapy appears to enhance and consolidate the antidepressant effects of esketamine, esketamine may in turn accelerate psychotherapeutic response by inducing transient states of enhanced neuroplasticity and emotional learning that increase receptivity to therapeutic interventions — facilitating the revision of maladaptive cognitive patterns and the reconsolidation of emotionally salient memories under reduced defensive responding (Krystal et al., 2024; Duman et al., 2019; Wilkinson et al., 2017; Wilkinson et al., 2021). Within this framework, KAP may function not merely as adjunctive support, but as a context capable of directing and integrating the neuroplastic and experiential processes elicited by esketamine administration.

KAP encourages participants to recognize, accept, and observe their emotional experiences — whether challenging or pleasant — from a more detached perspective, a process that may sensitize patients to perceive improvements in mood, functionality, and self-worth earlier during treatment. Although self-perception is often negatively biased in depressed individuals (Beck, 2021), this reorientation toward experiential awareness and acceptance may partially counteract that bias, contributing to the earlier emergence of between-group differences on patient-reported relative to clinician-rated measures. Although the present study was not designed to directly investigate these mechanisms, the delayed emergence of superior outcomes in the KAP condition appears compatible with models proposing an interaction between acute neurobiological effects and subsequent psychotherapeutic processing over time.

In light of these findings, an important clinical question emerges: if the adjunctive effects of psychotherapy take several weeks to become apparent, would it be reasonable to delay its introduction — reserving the initial sessions for the neurobiological action of esketamine alone, and reducing treatment complexity and associated costs in the process? We argue against this approach on several grounds. First, from a preparatory standpoint, psychotherapeutic support before the first administration serves functions that cannot be deferred: reducing treatment-related anxiety, providing psychoeducation about the effects of esketamine, establishing therapeutic agreements, and beginning the construction of the therapeutic alliance that will sustain patient engagement throughout treatment. Initiating psychotherapy only after pharmacological stabilization would deprive patients of this foundational preparation precisely at the moment when it is most needed — during their first encounter with dissociative and altered states of consciousness. Second, from an ethical perspective, as the field of psychedelic medicine continues to expand (Palhano-Fontes et al. 2019; Falchi-Carvalho et al. 2025), clinicians and researchers have increasingly argued that psychotherapeutic support represents not only a technical intervention but also an ethical component of patient care (Luoma et al., 2019; Nutt et al., 2025). Dissociative effects — including changes in sensory perception, thought, and identity — frequently occur with ketamine administration and may generate significant distress when unaccompanied by adequate psychological support (Mallevays et al., 2026). Third, from a temporal standpoint, evidence-based psychotherapies typically require four to eight weeks to produce clinically significant effects even under standard conditions (American Psychological Association, 2006; Beck, 2021; Leichsenring and Steinert, 2017); delaying its introduction would compress an already limited therapeutic window, potentially preventing the adjunctive effects observed in the present study from emerging at all. Early psychotherapeutic support may therefore contribute to establishing the relational and psychological conditions — safety, alliance, and openness — without which the integration of ketamine-induced neuroplastic processes into lasting behavioural and emotional change may not be possible. In this context, the early establishment of a therapeutic alliance may also facilitate treatment adherence, emotional safety, and comfort throughout the protocol — particularly during dissociative and altered states of consciousness that can be distressing when unaccompanied. Consistent with this interpretation, most participants in the KAP group reported feeling better supported in managing dissociative experiences and safer throughout treatment when accompanied by therapists responsible for preparing, supporting, and integrating their altered-state experiences.

The field of ketamine-based interventions encompasses a broad spectrum of psychological support models, ranging from purely pharmacological administration to fully structured psychotherapeutic protocols. At one end of this spectrum, ketamine is administered without any psychological support, with clinical outcomes attributed solely to its neurobiological effects (Zarate et al., 2006; Murrough et al., 2013). An intermediate model involves brief psychological support — typically one or two sessions focused on preparation or integration, without therapeutic accompaniment during administration — as exemplified by studies combining ketamine with motivational enhancement therapy for alcohol dependence (Grabski et al., 2022) or brief cognitive-behavioural interventions delivered before or after infusions (Wilkinson et al., 2021). At the other end of the spectrum lies fully integrated KAP, in which structured psychotherapeutic support encompasses all three phases of the therapeutic process — preparation, dosing accompaniment, and post-session integration — with explicit attention to psychoeducation, the therapeutic alliance, and the processing of dissociative and altered-state experiences, as employed in the present study. This gradation is clinically meaningful: the intensity, structure, and timing of psychological support vary substantially across studies, making direct comparisons of outcomes difficult and underscoring the need for standardized protocols that explicitly define the nature and scope of the psychotherapeutic component (Drozdz et al., 2022; Varela et al., 2026).

As previously cited, Moore et al. (2025) represents the only published study with objectives most closely aligned with those of the present investigation. Critically, the psychotherapy condition in that study did not include therapeutic support during ketamine administration — placing it within the intermediate model described above and representing a fundamental difference from the present protocol in which dosing accompaniment is considered both clinically and ethically essential (Luoma et al., 2022). Beyond this central distinction, several additional methodological differences may help explain the apparent discrepancy in outcomes. The study employed a naturalistic observational design with help-seeking samples of patients with depression and/or PTSD — a heterogeneous population that differs substantially from the present sample of patients with TRD. Although ketamine has shown therapeutic potential across both conditions (Pradhan et al., 2017; Velit-Salazar, Shiroma & Cherian, 2024), they involve distinct psychopathological mechanisms, and delivering effective psychotherapeutic interventions for two different diagnoses simultaneously within a brief treatment period represents a particular challenge. The treatment period was also shorter than that employed in the present study, and the psychotherapeutic protocol was not specifically developed or validated for either condition — in contrast to the present study, in which both the pharmacological and psychotherapeutic components were designed specifically for TRD. The authors themselves acknowledged that psychotherapeutic effects may not yet have become apparent within their 30-day follow-up window. Finally, participants in both groups could have received external psychotherapy without informing the research team, potentially attenuating between-group differences — a confound explicitly controlled for in the present study. Notably, exploratory analyses suggested that younger females showed better outcomes with combined treatment while older males showed better outcomes with ketamine alone, pointing to potential demographic moderators of treatment response that warrant further investigation.

Renal and hepatic function parameters remained stable throughout the treatment protocol in both groups, consistent with findings previously reported for Phase 1 of this trial (Palhano-Fontes et al., 2024) and with the broader literature on therapeutic ketamine use at subanesthetic doses (Kerr-Gaffney et al., 2025). Although recreational ketamine use at high doses and frequencies has been associated with urological toxicity — including ketamine-induced cystitis, bladder dysfunction, and upper urinary tract injury — clinical trials and observational studies using therapeutic ketamine for psychiatric disorders have reported urological symptoms in 0–24.5% of patients, with symptoms tending to be mild to moderate in severity and without significant changes in urinary parameters from baseline to follow-up (Kerr-Gaffney et al., 2025). Hepatobiliary abnormalities have also been reported in the context of recreational use, though evidence for hepatotoxicity with therapeutic doses remains limited and generally confined to high-dose sedation protocols rather than subanesthetic antidepressant regimens (Yoo et al., 2024). These distinctions are clinically important: the doses, frequencies, and duration employed in therapeutic protocols for TRD differ substantially from patterns associated with recreational toxicity and continued systematic monitoring of renal and urological function in long-term ketamine treatment studies remains warranted.

Both groups showed transient increases in blood pressure and heart rate following esketamine administration, with all parameters returning to baseline levels by the end of the session — confirming the cardiovascular safety profile of the subcutaneous esketamine protocol, consistent with Phase 1 findings (Palhano-Fontes et al., 2024) and with the broader literature on subanesthetic ketamine administration (Loo et al., 2023). No adverse cardiovascular events were recorded in either group throughout the study.

Notably, the KAP group showed earlier return to baseline in both systolic and diastolic blood pressure, and significantly lower diastolic values compared to the KET group from +30 to +60 minutes. This pattern may reflect an attenuation of the sympathetic cardiovascular response associated with the psychotherapeutic context — including the therapeutic alliance, preparatory psychoeducation, and the presence of trained therapists during dosing — which may have contributed to greater emotional safety and reduced physiological stress reactivity during the session (Porges, 2011).

An intriguing finding concerns heart rate: while the KET group showed a progressive decline from baseline throughout the session, the KAP group exhibited a transient increase at +15 minutes followed by a progressive reduction below baseline values at later timepoints. This divergent pattern may tentatively suggest that the psychotherapeutic context — by encouraging openness and emotional engagement with the altered-state experience — elicits an initial autonomic response reflecting increased emotional processing, followed by a deeper parasympathetic relaxation response over the course of the session. A similar interpretive framework may apply to the divergent oxygen saturation patterns observed between groups. However, directly testing these hypotheses would require cross-referencing cardiovascular data with measures of dissociative state intensity — such as CADSS scores — which falls outside the primary scope of the present investigation and will be examined in subsequent analyses.

Some limitations of the present study should be acknowledged. First, the non-randomized design — inherent to the sequential phase structure — represents a methodological constraint, despite the comparable demographic and clinical characteristics observed between groups at baseline. Second, no placebo condition was employed for either esketamine administration or psychotherapy; comparisons were therefore restricted to two groups receiving active interventions, and the specific contribution of expectancy effects cannot be fully excluded. In this context, it is possible that the perception of increased psychosocial support and access to a broader multidisciplinary care network may have contributed to mood improvements in the KAP group, including beyond the formal treatment period. Third, the therapeutic setting differed between conditions: esketamine administrations in the KET group were conducted in a standard hospital outpatient environment, whereas KAP sessions took place in an individualized, non-clinical therapeutic setting. While this difference should be considered when interpreting the findings, it is important to note that the setting is not merely an incidental variable — it is an integral component of each treatment model. Replicating the KAP environment for the KET condition, or vice versa, would compromise the ecological validity and clinical authenticity of at least one of the protocols. In this sense, the distinction in setting reflects the real-world differences between biomedical and psychotherapy-integrated approaches to ketamine treatment, and its potential contribution to outcomes warrants explicit investigation in future studies. Fourth, clinician-rated depressive symptom evaluation using the MADRS was not continued beyond the end of treatment, with follow-up relying exclusively on self-report measures — precluding a more rigorous examination of whether the adjunctive effects of KAP were sustained, attenuated, or reversed in the months following protocol completion. Whether participants continued psychotherapy after study completion was also not systematically assessed, representing an uncontrolled variable that may have differentially influenced long-term outcomes between groups. Finally, the relatively small sample size limits the statistical power and generalizability of the findings. Future studies investigating the adjunctive role of psychotherapy in ketamine-based interventions should incorporate clinician-rated assessments throughout follow-up, control for the continuity of psychotherapeutic engagement after treatment, and seek replication in larger, randomized controlled trials.

## Conclusion

This study provides the first clinical evidence that structured ketamine-assisted psychotherapy — encompassing preparation, dosing accompaniment, and post-session integration — participants receiving esketamine within the KAP model showed superior end-of-treatment outcomes in patients with TRD, with notably higher response and remission rates and earlier subjective symptom improvement in the KAP condition. These findings are consistent with the possibility that the acute psychoactive experience elicited by esketamine is not merely a pharmacological byproduct to be managed, but an active therapeutic component that can be meaningfully shaped by structured psychological support. The cardiovascular safety profile was favourable in both conditions, with all parameters returning to baseline by session end, and the KAP group showing earlier hemodynamic normalization — consistent with a reduced sympathetic stress response in the context of therapeutic accompaniment. Together, these results support further investigation in which esketamine-induced neuroplasticity and psychotherapeutic processing may act synergistically to enhance clinical outcomes in TRD.

Several challenges remain. The absence of randomization, placebo control, and clinician-rated follow-up assessments limits the conclusions that can be drawn, and the small sample size warrants caution in generalizing these findings. Future studies should employ more rigorous designs, incorporate structured and theoretically defined psychotherapeutic approaches, continue clinician-rated assessment through follow-up, and investigate the mechanisms underlying the interaction between neurobiological and psychological components of KAP — including the role of dissociative states, therapeutic alliance, and post-session integration in determining both short- and long-term outcomes.

## Data Availability

All data produced in the present study are available upon reasonable request to the authors
Some data produced in the present work are contained in the manuscript

## FUNDING

This research received no specific grant from any funding agency in the public, commercial, or not-for-profit sectors.

## Supplementary material

**Figure S1.**
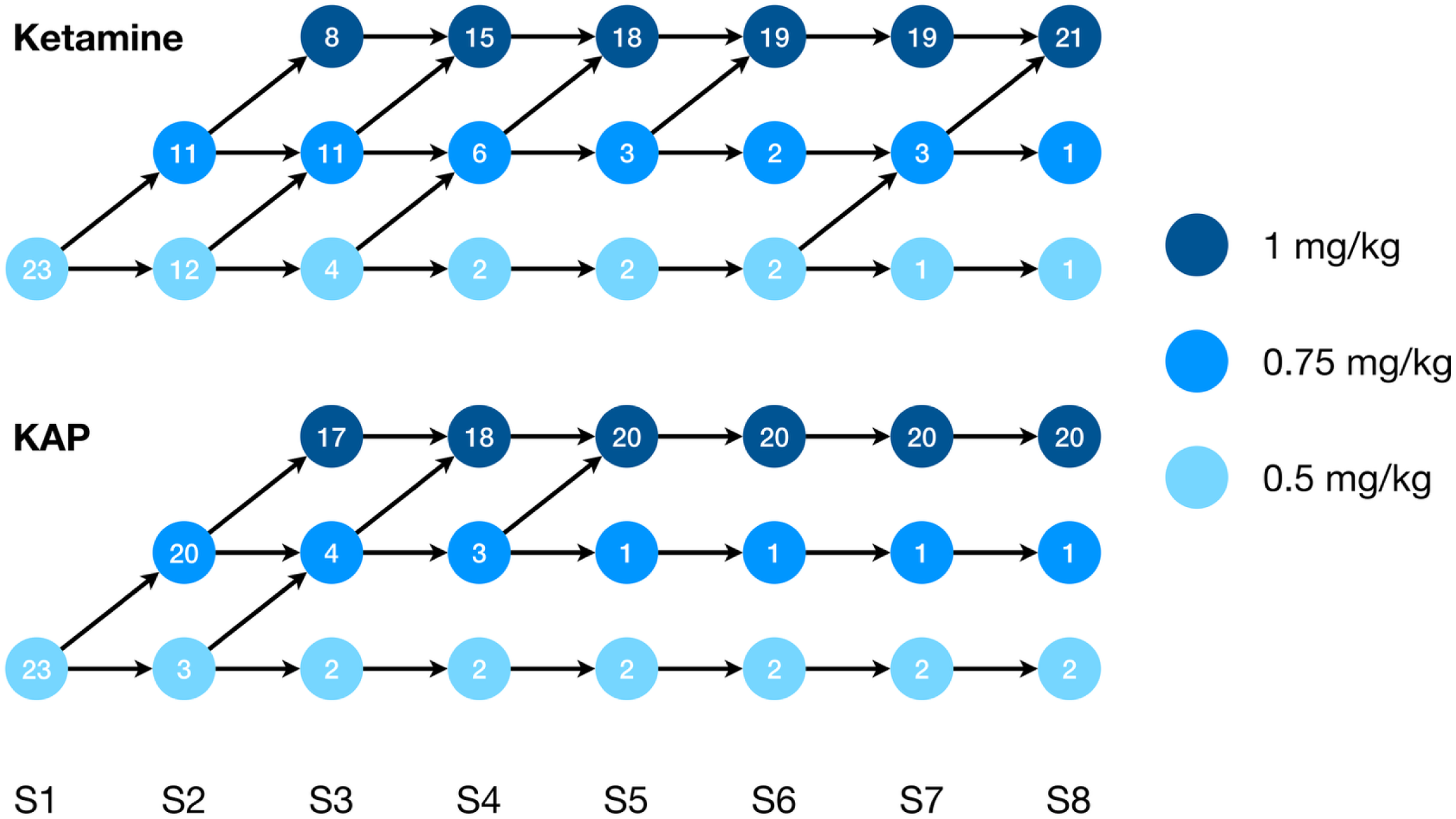
Dose distribution through the sessions. All patients received 0.5 mg/kg at the first dosing session. The doses were increased by 0.25 mg/kg in the next dosing session up to 1 mg/kg if a clinical response was not met (reduction of at least 50% of the MADRS from the previous session or did not reach remission). The lowest dose at which we observed a response was repeated throughout treatment.

**Figure S2.**
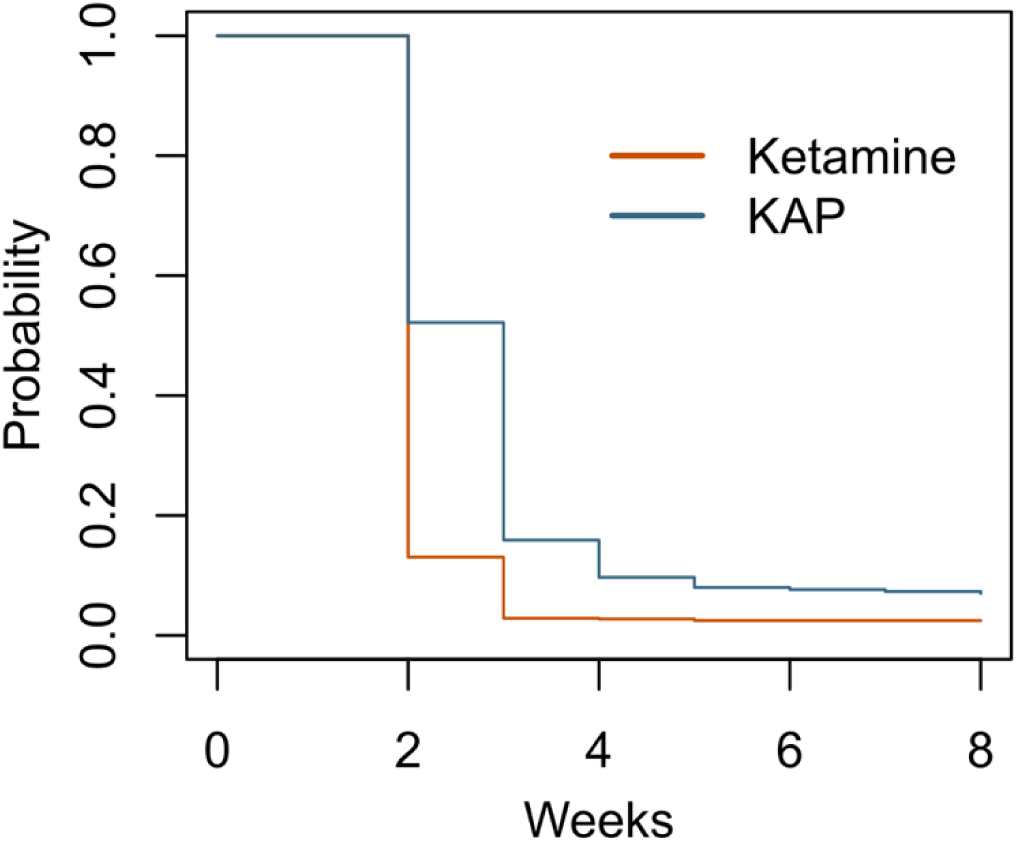
Estimated probability curves over an 8-week follow-up period for the Ketamine and Ketamine-Assisted Psychotherapy (KAP) treatment groups. The dose evolution was statistically similar for both groups (HR = 0.90; 95% CI: 0.67 – 1.22; p = 0.507).

**Table S1.** Biochemistry values for all time points and groups.

|  |  | Ketamine |  |  | KAT |  |  | Statistical analysis<br>(group*session) |
| --- | --- | --- | --- | --- | --- | --- | --- | --- |
|  | Reference range | Baseline | 4 weeks | 8 weeks | Baseline | 4 weeks | 8 weeks |  |
| AST | Men: up to 38 U/L<br>Women: up to 32 U/L | 24.91 (±11.64) | 24.83 (±13.38) | 29.04 (±22.89) | 20.85 (±5.97) | 23.36 (±12.94) | 24.99 (±13.78) | F (1.23, 49.19) = 0.208; p=0.701 |
| ALT | Men: up to 37 U/L<br>Women: up to 45 U/L | 29.35 (±21.19) | 29.87 (±22.81) | 34.48 (±40.85) | 22.59 (±12.76) | 31.31 (±34.35) | 25.19 (±22.64) | F (1.19, 45.04) = 0.926; p=0.357 |
| Urea | Men: 19 - 43 mg/dL<br>Women: 15 - 36 mg/dL | 32.22 (±11.15) | 31.61 (±9.63) | 30.96 (±8.22) | 33.09 (±9.48) | 34.39 (±10.73) | 32.32 (±9.34) | F (2, 82) = 0.169; p=0.845 |
| Creatinine | Men: 0.7 – 1.3 mg/dL<br>Women: 0.6 – 1.1 mg/dL | 0.94 (±0.26) | 0.88 (±0.22) | 0.88 (±0.21) | 0.87 (±0.18) | 0.90 (±0.23) | 0.86 (±0.21) | F (2, 80) = 2.397; p=0.097 |
| Na<sup>+</sup> | 135 – 146 mEq/L | 138.86 (±5.23) | 139.65 (±3.94) | 139.41 (±5.11) | 140.25 (±2.06) | 141.19 (±3.00) | 140.77 (±3.37) | F (2, 82) = 0.083; p=0.920 |
| K<sup>+</sup> | 3.5 – 5.0 mEq/L | 4.13 (±0.38) | 4.10 (±0.30) | 4.17 (±0.42) | 4.33 (±0.33) | 4.30 (±0.31) | 4.21 (±0.32) | F (2, 84) = 1.138; p=0.325 |

AST: Aspartate aminotransferase; ALT: Alanine aminotransferase

**Table S2.** Vital signals during the acute (baseline and minutes after infusion) esketamine effects for both groups. Values are mean (±standard deviation) throughout the eight sessions.

|  | Basal | +15 | +30 | +45 | +60 |
| --- | --- | --- | --- | --- | --- |
| ***Ketamine*** |  |  |  |  |  |
| Systolic Blood Pressure (mmHg) | 119.38 (±1.01) | 131.54 (±1.28) | 130.11 (±1.20) | 126.76 (±1.11) | 124.67 (±1.05) |
| Diastolic Blood Pressure (mmHg) | 79.17 (±0.79) | 87.17 (±0.79) | 86.49 (±0.78) | 84.27 (±0.78) | 83.86 (±0.77) |
| Heart Rate (bpm) | 80.69 (±1.05) | 81.16 (±1.05) | 78.05 (±1.04) | 76.71 (±0.98) | 76.35 (±0.96) |
| SpO<sub>2</sub> (%) | 97.15 (±0.14) | 97.60 (±0.14) | 97.55 (±0.14) | 97.60 (±0.15) | 97.68 (±0.16) |
| ***KAP*** |  |  |  |  |  |
| Systolic Blood Pressure (mmHg) | 122.83 ($\pm 0.97$) | 134.34 ($\pm 1.23$) | 132.99 ($\pm 1.16$) | 129.29 ($\pm 1.07$) | 125.07 ($\pm 1.01$) |
| Diastolic Blood Pressure (mmHg) | 75.83 ($\pm 0.76$) | 83.93 ($\pm 0.76$) | 82.81 ($\pm 0.75$) | 79.59 ($\pm 0.75$) | 77.59 ($\pm 0.74$) |
| Heart Rate (bpm) | 76.32 ($\pm 1.01$) | 82.47 ($\pm 1.01$) | 80.45 ($\pm 1.00$) | 76.62 ($\pm 0.94$) | 75.31 ($\pm 0.92$) |
| SpO<sub>2</sub> (%) | 97.93 ($\pm 0.13$) | 97.98 ($\pm 0.13$) | 97.85 ($\pm 0.13$) | 97.47 ($\pm 0.14$) | 97.23 ($\pm 0.15$) |

**Table S3.** Pairwise comparisons (mean difference and adjusted p-values) for vital signals between and within groups.

| Between-groups | SBP | DBP | HR | SpO<sub>2</sub> |
| --- | --- | --- | --- | --- |
| 0 | -3.46 (0.290) | 3.34 (0.073) | 4.37 (0.084) | 0.78 (0.002) |
| +15 | -2.80 (0.860) | 3.24 (0.093) | -1.31 (0.996) | 0.39 (0.619) |
| +30 | -2.88 (0.778) | 3.67 (0.025) | -2.40 (0.815) | 0.30 (0.850) |
| +45 | -2.53 (0.828) | 4.68 (<0.001) | 0.08 (1.000) | -0.13 (1.000) |
| +60 | -0.400 (1.000) | 6.27 (<0.001) | 1.04 (0.999) | -0.51 (0.552) |
| Within-group | Ketamine |  | KAP |  |
|  | Mean difference | Adjusted p-value | Mean difference | Adjusted p-value |
| *Systolic blood pressure* |  |  |  |  |
| 0 vs. +15 | -12.17 | <0.001 | -11.50 | <0.001 |
| 0 vs. +30 | -10.74 | <0.001 | -10.16 | <0.001 |
| 0 vs. +45 | -7.39 | <0.001 | -6.45 | <0.001 |
| 0 vs. +60 | -5.29 | <0.001 | -2.23 | 0.306 |
| *Diastolic blood pressure* |  |  |  |  |
| 0 vs. +15 | -8.00 | <0.001 | -8.10 | <0.001 |
| 0 vs. +30 | -7.33 | <0.001 | -6.98 | <0.001 |
| 0 vs. +45 | -5.11 | <0.001 | -3.76 | <0.001 |
| 0 vs. +60 | -4.69 | <0.001 | -1.76 | 0.277 |
| **Heart rate** |  |  |  |  |
| 0 vs. +15 | -0.47 | 1.000 | -6.15 | <0.001 |
| 0 vs. +30 | 2.64 | 0.036 | -4.13 | <0.001 |
| 0 vs. +45 | 3.98 | <0.001 | -0.304 | 1.000 |
| 0 vs. +60 | 4.33 | <0.001 | 1.01 | 0.895 |
| **SpO<sub>2</sub>** |  |  |  |  |
| 0 vs. +15 | -0.45 | 0.027 | -0.06 | 1.000 |
| 0 vs. +30 | -0.40 | 0.062 | 0.08 | 1.000 |
| 0 vs. +45 | -0.45 | 0.047 | 0.46 | 0.019 |
| 0 vs. +60 | -0.53 | 0.012 | 0.69 | <0.001 |

**Table S4.** Response and remission rates across sessions.

|  | Response |  |  | Remission |  |  |
| --- | --- | --- | --- | --- | --- | --- |
| | Ketamine | KAP | $\chi^2$ (p-value) | Ketamine | KAP | $\chi^2$ (p-value) |
| **S1** | 2 (8.7%) | 3 (13.0%) | 0.00 (1.000) | 1 (4.3%) | 2 (8.7%) | 0.00 (1.000) |
| **S2** | 3 (13%) | 2 (8.7%) | 0.00 (1.000) | 3 (13%) | 3 (13.0%) | 0.00 (1.000) |
| **S3** | 7 (30.4%) | 8 (34.8%) | 0.00 (1.000) | 6 (26.1%) | 6 (26.1%) | 0.00 (1.000) |
| **S4** | 9 (39.1%) | 9 (39.1%) | 0.00 (1.000) | 6 (26.1%) | 6 (26.1%) | 0.00 (1.000) |
| **S5** | 10 (43.5%) | 12 (52.2%) | 0.09 (0.768) | 9 (39.1%) | 12 (52.2%) | 0.35 (0.554) |
| **S6** | 12 (52.2%) | 13 (56.5%) | 0.00 (1.000) | 9 (39.1%) | 13 (56.5%) | 0.78 (0.376) |
| **S7** | 12 (52.2%) | 14 (60.9%) | 0.09 (0.767) | 9 (39.1%) | 14 (60.9%) | 1.39 (0.238) |
| **S8** | 12 (52.2%) | 18 (78.3%) | 2.40 (0.122) | 8 (34.8%) | 18 (78.3%) | 7.17 (0.007) |

**Table S5.** Within-group effect size and p-value of the comparison regarding baseline.

|  | Ketamine |  | KAP |  |
| --- | --- | --- | --- | --- |
|  | d (95% IC) | p-value | d (95% IC) | p-value |
| **MADRS** |  |  |  |  |
| Session 1 | -0.757 (-1.118; -0.395) | 0.004 | -0.721 (-1.076; -0.366) | 0.014 |
| Session 2 | -1.271 (-1.734; -0.809) | < 0.001 | -0.872 (-1.254; -0.491) | 0.002 |
| Session 3 | -1.463 (-1.969; -0.957) | < 0.001 | -1.499 (-2.014; -0.985) | < 0.001 |
| Session 4 | -1.338 (-1.815; -0.86) | < 0.001 | -1.743 (-2.316; -1.17) | < 0.001 |
| Session 5 | -1.487 (-1.998; -0.976) | < 0.001 | -1.704 (-2.267; -1.141) | < 0.001 |
| Session 6 | -1.377 (-1.863; -0.891) | < 0.001 | -2.275 (-2.982; -1.569) | < 0.001 |
| Session 7 | -1.267 (-1.729; -0.806) | < 0.001 | -2.597 (-3.387; -1.807) | < 0.001 |
| Session 8 | -1.959 (-2.585; -1.333) | < 0.001 | -2.476 (-3.234; -1.717) | < 0.001 |

| **BDI – D1** |  |  |  |  |
| --- | --- | --- | --- | --- |
| Session 1 | -0.651 (-0.995; -0.306) | 0.018 | -1.037 (-1.450; -0.624) | < 0.001 |
| Session 2 | -0.729 (-1.085; -0.372) | 0.007 | -1.276 (-1.739; -0.812) | < 0.001 |
| Session 3 | -0.654 (-0.998; -0.309) | 0.012 | -1.417 (-1.912; -0.922) | < 0.001 |
| Session 4 | -0.928 (-1.319; -0.536) | < 0.001 | -1.436 (-1.935; -0.936) | < 0.001 |
| Session 5 | -1.227 (-1.68; -0.774) | < 0.001 | -1.604 (-2.143; -1.065) | < 0.001 |
| Session 6 | -1.102 (-1.529; -0.676) | < 0.001 | -1.573 (-2.104; -1.041) | < 0.001 |
| Session 7 | -1.184 (-1.628; -0.741) | < 0.001 | -2.00 (-2.636; -1.364) | < 0.001 |
| Session 8 | -1.327 (-1.802; -0.852) | < 0.001 | -1.784 (-2.367; -1.202) | < 0.001 |
| **BDI – D7** |  |  |  |  |
| Session 1 | -0.433 (-0.748; -0.117) | 0.115 | -0.848 (-1.225; -0.471) | 0.004 |
| Session 2 | -0.700 (-1.052; -0.348) | 0.011 | -1.123 (-1.553; -0.692) | < 0.001 |
| Session 3 | -0.689 (-1.039; -0.339) | 0.010 | -1.327 (-1.802; -0.852) | < 0.001 |
| Session 4 | -0.703 (-1.056; -0.351) | 0.008 | -1.584 (-2.119; -1.050) | < 0.001 |
| Session 5 | -1.075 (-1.496; -0.654) | < 0.001 | -1.445 (-1.947; -0.943) | < 0.001 |
| Session 6 | -1.118 (-1.548; -0.688) | < 0.001 | -1.777 (-2.357; -1.196) | < 0.001 |
| Session 7 | -1.067 (-1.486; -0.648) | < 0.001 | -1.699 (-2.261; -1.137) | < 0.001 |
| Session 8 | -1.319 (-1.792; -0.846) | < 0.001 | -1.861 (-2.462; -1.259) | < 0.001 |
| **MADRS-SI** |  |  |  |  |
| Session 1 | -0.441 (-0.874; -0.008) | 0.133 | -0.727 (-1.106; -0.349) | 0.014 |
| Session 2 | -0.736 (-1.203; -0.269) | 0.011 | -0.558 (-1.036; -0.08) | 0.063 |
| Session 3 | -0.671 (-1.085; -0.257) | 0.022 | -1.123 (-1.679; -0.567) | < 0.001 |
| Session 4 | -0.646 (-1.138; -0.154) | 0.028 | -1.477 (-2.102; -0.852) | < 0.001 |
| Session 5 | -0.736 (-1.149; -0.324) | 0.012 | -1.45 (-2.062; -0.837) | < 0.001 |
| Session 6 | -0.855 (-1.338; -0.372) | 0.004 | -1.489 (-2.135; -0.843) | < 0.001 |
| Session 7 | -0.370 (-0.722; -0.018) | 0.247 | -1.588 (-2.257; -0.92) | < 0.001 |
| Session 8 | -1.201 (-1.781; -0.622) | < 0.001 | -1.537 (-2.193; -0.882) | < 0.001 |

**Table S6.** Estimated marginal means (and standard error), effect size, and p-value for the between-group comparisons across outcomes.

|  | **Ketamine** | **KAP** | **d (95% IC)** | **p-value** |
| --- | --- | --- | --- | --- |
| **MADRS** |  |  |  |  |
| Basal | 30.93 (0.9995) | 28.37 (1.2623) | 0.469 (-0.033; 0.971) | 0.111 |
| Session 1 | 25.63 (1.5775) | 23.55 (1.4778) | 0.284 (-0.214; 0.782) | 0.335 |
| Session 2 | 22.28 (1.5355) | 22.02 (1.6388) | 0.034 (-0.461; 0.53) | 0.909 |
| Session 3 | 19.32 (1.7643) | 17.29 (1.6658) | 0.247 (-0.251; 0.744) | 0.401 |
| Session 4 | 19.63 (1.8625) | 15.68 (1.6387) | 0.470 (-0.032; 0.972) | 0.111 |
| Session 5 | 16.89 (2.0473) | 15.24 (1.7421) | 0.181 (-0.315; 0.677) | 0.541 |
| Session 6 | 16.80 (2.1952) | 13.07 (1.4894) | 0.415 (-0.086; 0.915) | 0.159 |
| Session 7 | 17.63 (2.2368) | 11.07 (1.4712) | 0.723 (0.212; 1.233) | 0.015 |
| Session 8 | 15.15 (1.7875) | 10.15 (1.6582) | 0.605 (0.098; 1.111) | 0.041 |
| **BDI – D1** |  |  |  |  |
| Basal | 37.905 (1.9117) | 35.442 (2.3579) | 0.239 (-0.258; 0.736) | 0.420 |
| Session 1 | 30.079 (2.7205) | 23.355 (2.4939) | 0.537 (0.033; 1.041) | 0.069 |
| Session 2 | 28.645 (2.8704) | 20.268 (2.5757) | 0.641 (0.133; 1.148) | 0.030 |
| Session 3 | 28.427 (3.2405) | 19.529 (2.3245) | 0.658 (0.150; 1.166) | 0.026 |
| Session 4 | 25.427 (3.0281) | 18.747 (2.4823) | 0.503 (0.000; 1.006) | 0.089 |
| Session 5 | 23.645 (2.6270) | 17.095 (2.4107) | 0.542 (0.038; 1.046) | 0.066 |
| Session 6 | 22.123 (3.2045) | 16.486 (2.6275) | 0.401 (-0.099; 0.901) | 0.174 |
| Session 7 | 21.210 (3.1600) | 13.790 (2.1219) | 0.575 (0.070; 1.080) | 0.052 |
| Session 8 | 21.514 (2.7922) | 14.790 (2.4625) | 0.532 (0.028; 1.036) | 0.071 |

| **BDI – D7** |  |  |  |  |
| --- | --- | --- | --- | --- |
| Basal | 37.88 (1.9013) | 35.47 (2.3615) | 0.234 (-0.263; 0.732) | 0.429 |
| Session 1 | 32.71 (2.7031) | 25.69 (2.4442) | 0.568 (0.063; 1.073) | 0.055 |
| Session 2 | 29.45 (2.7246) | 23.08 (2.2304) | 0.533 (0.030; 1.037) | 0.071 |
| Session 3 | 28.84 (2.9586) | 20.69 (2.2780) | 0.644 (0.136; 1.151) | 0.029 |
| Session 4 | 28.36 (3.0447) | 17.95 (2.2417) | 0.812 (0.297; 1.327) | 0.006 |
| Session 5 | 23.23 (3.0629) | 18.25 (2.5807) | 0.367 (-0.133; 0.866) | 0.215 |
| Session 6 | 23.58 (2.8878) | 15.95 (2.2057) | 0.619 (0.112; 1.126) | 0.036 |
| Session 7 | 24.58 (2.8181) | 14.82 (2.4276) | 0.774 (0.261; 1.287) | 0.009 |
| Session 8 | 20.88 (2.9086) | 13.77 (2.4920) | 0.547 (0.043; 1.052) | 0.063 |
| **MADRS-SI** |  |  |  |  |
| Basal | 2.514 (0.3325) | 1.824 (0.2209) | -1.537 (-2.193; -0.882) | 0.085 |
| Session 1 | 1.818 (0.3253) | 1.052 (0.2260) | 0.511 (0.002; 1.02) | 0.054 |
| Session 2 | 1.470 (0.2442) | 1.233 (0.2306) | 0.570 (0.065; 1.075) | 0.482 |
| Session 3 | 1.470 (0.3157) | 0.704 (0.1993) | 0.210 (-0.293; 0.713) | 0.041 |
| Session 4 | 1.470 (0.3416) | 0.486 (0.1508) | 0.605 (0.099; 1.111) | 0.008 |
| Session 5 | 1.297 (0.3555) | 0.486 (0.1607) | 0.777 (0.264; 1.290) | 0.038 |
| Session 6 | 1.265 (0.2751) | 0.486 (0.1490) | 0.613 (0.106; 1.120) | 0.013 |
| Session 7 | 1.870 (0.4482) | 0.400 (0.1480) | 0.752 (0.233; 1.270) | 0.002 |
| Session 8 | 0.895 (0.2995) | 0.443 (0.1488) | 1.118 (0.540; 1.695) | 0.178 |

**Table S7.**
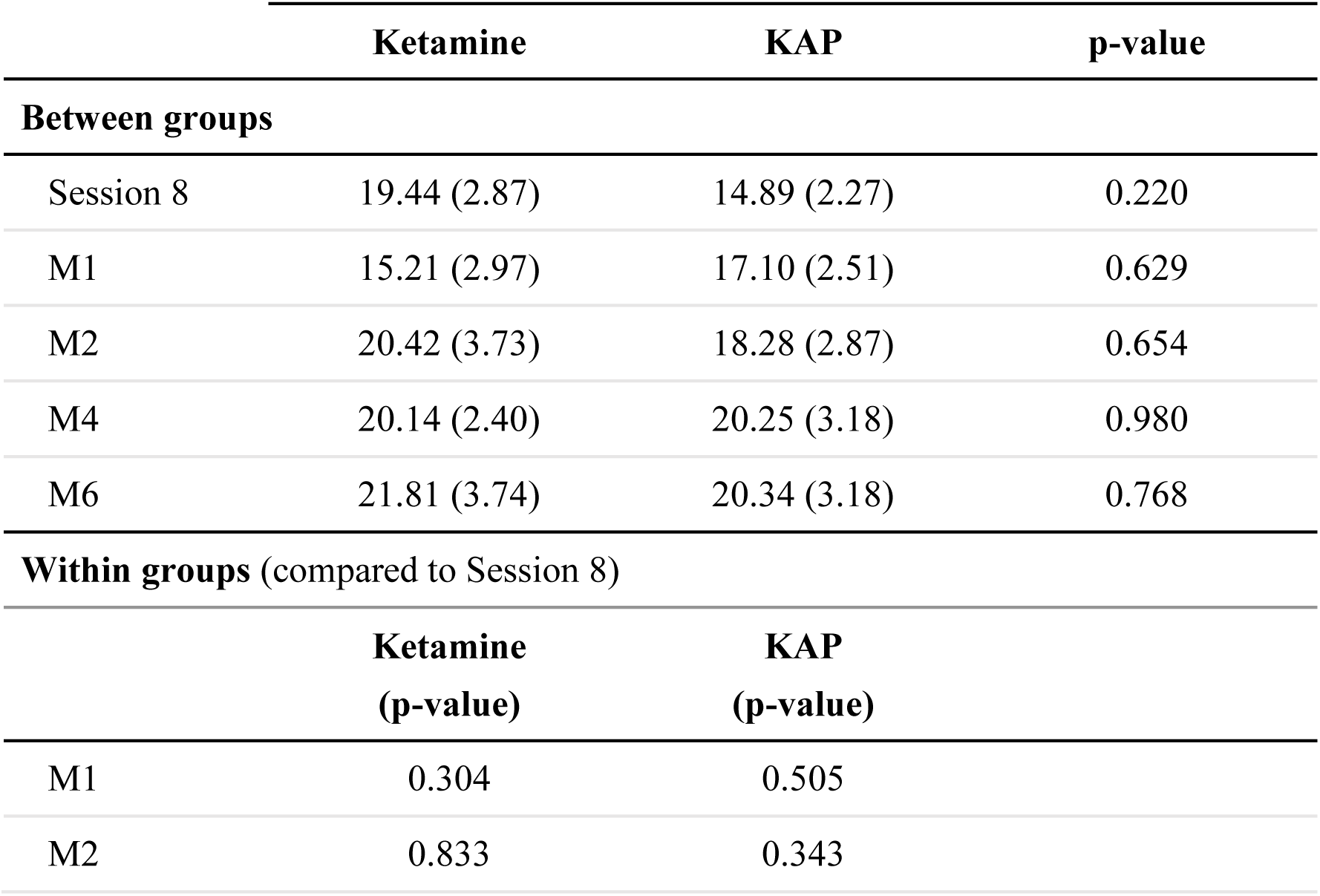
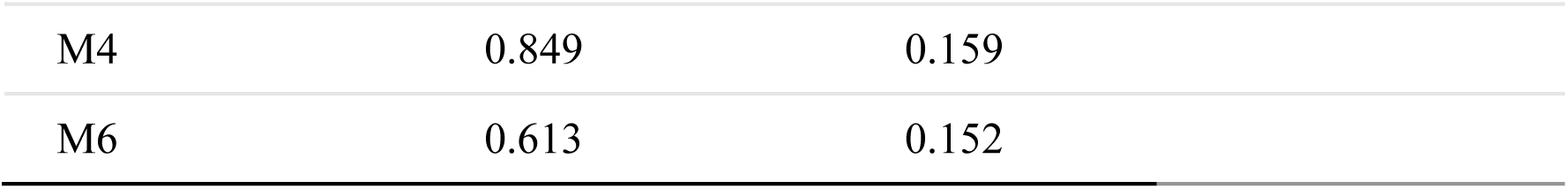
Marginal means and comparisons within and between groups for the BDI follow-up.

|  | **Ketamine** | **KAP** | **p-value** |
| --- | --- | --- | --- |
| **Between groups** |  |  |  |
| Session 8 | 19.44 (2.87) | 14.89 (2.27) | 0.220 |
| M1 | 15.21 (2.97) | 17.10 (2.51) | 0.629 |
| M2 | 20.42 (3.73) | 18.28 (2.87) | 0.654 |
| M4 | 20.14 (2.40) | 20.25 (3.18) | 0.980 |
| M6 | 21.81 (3.74) | 20.34 (3.18) | 0.768 |
| **Within groups (compared to Session 8)** |  |  |  |
|  | **Ketamine**<br>**(p-value)** | **KAP**<br>**(p-value)** |  |
| M1 | 0.304 | 0.505 |  |
| M2 | 0.833 | 0.343 |  |

|  |  |  |
| --- | --- | --- |
| M4 | 0.849 | 0.159 |
| M6 | 0.613 | 0.152 |

## Notes

### Competing Interest Statement

The authors have declared no competing interest.

### Clinical Trial

The trial was registered at https://ensaiosclinicos.gov.br/rg/RBR-1072m6nv

### Author Declarations

This study was conducted at the Hospital Universitário Onofre Lopes (HUOL/UFRN) and approved by the institution's Research Ethics Committee under protocol number 39222920.2.0000.5292

